# The development and validation of a menu healthiness assessment tool (Menu-Health) for small-to-medium sized businesses in the UK out-of-home food sector

**DOI:** 10.1101/2025.11.10.25339876

**Authors:** Jane Brealey, Joanna Brooks, Zoé Colombet, Amy Finlay, Eric Robinson

**Author notes:** Funder: This work was funded by the Economic and Social Research Council.

## Abstract

**Background:** Out-of-home eating makes a significant contribution to dietary intake. Food consumed out- of-home is often higher in energy and less nutritious than food consumed at home. Evaluating the healthiness of catering practices in the out-of-home food sector (OHFS) is challenging for small-to-medium sized businesses, and validated assessment tools are limited. The objective of the present research was to develop and validate a menu healthiness assessment tool for small-to-medium sized UK OHFS businesses.

**Methods:** We devised and validated a menu healthiness assessment tool, Menu-Health, based on UK nutritional guidance and recommendations. Content validity was assessed by six independent experts. Inter-rater reliability and predictive validity were assessed by applying the tool to 66 OHFS businesses in the Northwest of England. We sampled 33 businesses with an accredited tiered local authority healthier catering award. These businesses were paired with similar non-award holder businesses, matched by business type, cuisine type and Indices of Multiple Deprivation (IMD) decile.

**Results:** Inter-rater agreement for overall menu healthiness assessment score was 92%. The tool also had strong inter-rater agreement and excellent inter-rater reliability. Only 18% of assessed OHFS businesses had a ‘good’ overall healthiness score (≥71%). Regression analyses examined if award status or award level, business type and IMD decile predicted healthiness scores. As expected, takeaways and dessert outlets had lower healthiness scores than cafés, restaurants and pubs, as did outlets located in more deprived areas (IMD 1-2 vs. 3-10). Award holder status was not significantly associated with healthiness scores. However, higher award tier level was associated with higher menu healthiness scores.

**Conclusion:** Menu-Health provides a comprehensive, validated and reliable method of assessing and comparing the food, drink and catering practices of small-to-medium sized UK OHFS businesses. It is potentially a useful tool for researchers, public health teams and local authorities to evaluate the healthiness of local OHFS environments.

## 1. Introduction

Trends for consuming food from the out-of-home food sector (OHFS^1^) have increased both in Europe [1, 2] and globally [3, 4], resulting in the OHFS making a substantial contribution to diet. A US Department of Agriculture analysis estimated the OHFS accounted for >30% of US food energy intake in 2018 [5], compared to 18% in 1977-1978 [6]. In the UK, between 11-15% of the general population’s daily energy intake is from the OHFS [7]. Data from the UK National Diet and Nutrition Survey (NDNS 2019-2023) show that 72% of people in England had purchased food or drink from the OHFS in the seven days before the study [8].

Research in the UK found that food sold in independent takeaways contained excessive energy and salt, and was large in portion size [9, 10]. A US study found that food sold in independent OHFS businesses contained more energy than food sold in larger chain restaurants [11]. Concerns have also been raised globally about the nutritional quality of food in the OHFS; US and Canadian research shows that food purchased out-of-home contains higher amounts of saturated fat, sodium and refined grains, and limited amounts of fibre, wholegrains, fruit, dairy and vegetables [5, 12–14].

In an attempt to improve the healthiness of the out-of-home food environment, and thus tackle the obesity burden, the UK government implemented salt, sugar and calorie reduction targets between 2017 and 2022 [15, 16]. A mandatory calorie labelling policy was also introduced for large businesses in 2022 to ‘help people make healthier choices’ [17–19].

Although small-medium sized businesses (SMEs^2^) were encouraged to adopt the policy, they were not legally mandated to do so [17]. Yet SMEs make a large contribution to the UK diet; in 2022, 82% of OHFS outlets in the UK were classed as SMEs, accounting for 53% of the income in the OHFS [20].

Alongside central government action, local government plays an important role in improving the health of local food environments through public health and planning interventions [21–23]. For example, in the UK, some local governments use healthier catering awards or accreditation schemes [22] to reward healthy menu practices and incentivise SMEs to improve the healthiness of food and drink sold. Such schemes have been introduced globally in OHFS businesses, grocery stores, workplace cafeterias, schools, childcare settings, vending machines and hospitals. Schemes vary, but to receive accreditation, businesses are typically required to add healthier options to menus, such as those with less fat or less salt or those with more fruit, vegetables and wholegrains [21, 24–30], offer smaller portions [24, 28], and label/promote healthier options [28, 31, 32].

Findings of individual schemes’ impact and success are mixed, and evidence varies in quality [33–35]. Evaluations are commonly limited because they examine compliance to scheme criteria, rather than overall healthiness of food, drink and catering practices, For example, an analysis of the ‘Shape up Somerville’ initiative (Massachusetts, US) assessed impact by examining a set of criteria specific to the scheme, as opposed to using a validated menu healthiness assessment tool [28]. A similar analysis of the ‘Choose Health LA’ scheme (Los Angeles, US) examined menu changes post scheme implementation, again focusing on scheme criteria rather than overall menu healthiness [24]. Similarly, a UK study analysing the Healthier Catering Commitment was unable to quantify overall impact of the scheme on business practice (i.e. healthiness of menus) and instead examined the number of changes businesses self-reported [21]. A major limitation of research to date is reliance on business reported compliance to menu healthiness principles, with limited validated and appropriate assessment tools for OHFS business menu healthiness [36, 37].

A small number of eating environment / menu assessment tools have been developed, but most have limited application to the UK OHFS. The Nutrition Environment Measures Study in Restaurants (NEMS-R) is one of the most commonly used observational food environment audit tools, validated and tested in a US context in 2004-2005 [38]. NEMS-R was found to have high levels of inter-rater percent agreement (>75%), and moderate inter-rater reliability scores (Kappa range 0.27-0.97, mean Kappa 0.69)[38]. Although NEMS-R has been adapted by a number of researchers to assess a variety of food environments [39], NEMS-R has been found to be labour and time intensive [40]. Research also suggests application to countries outside of the US may not provide reliable assessments of the food environment [41].

More recently, a desk-based Menu Assessment Scoring Tool (MAST) was developed using Australian dietary guidelines by researchers in Perth, Western Australia [42]. The tool used similar methodology to the Portuguese Kids’ Menu Healthy Score (KIMEHS) [43] and the Brazilian Healthiness Indicators scoring system [44], whereby a risk assessment approach was utilised to classify menu items as either nutritious or nutrient-poor, with penalty points applied for the latter. Researchers developing the MAST system found that assessing SMEs using a purely desk-based approach was beneficial from a time perspective, but collecting accurate data was challenging due to the absence of online menus or omission of menu items e.g. daily specials, seasonal variations and soft drinks. Furthermore, the tool does not capture data on cooking methods, portion sizes, prices and promotions, and validity and reliability were not formally assessed [42]. In 2020, UK-based researchers developed an exploratory menu healthiness scoring system for use in the online takeaway sector [45], but the measure did not assess the overall healthiness of menus or healthier catering practices.

The current lack of UK OHFS menu healthiness assessment tools is a challenge to evaluation of interventions designed to improve SME OHFS menu healthiness in the UK. Furthermore, the lack of a validated tool prevents monitoring of the healthiness of OHFS businesses (e.g. geographical trends) and changes over time. At present, only very large businesses report limited nutrition information (e.g. calorie content of menu items), and assessment of OHFS menu healthiness reliant on such information is not feasible in SMEs [46]. Because of this, there is a need to develop a menu healthiness assessment tool for the UK OHFS which does not rely on reported nutritional information and can be applied to SMEs.

No appropriate menu assessment tools exist for assessing OHFS menu healthiness in UK SMEs. Moreover, the small number of existing menu healthiness tools developed to date have limitations and have tended not to be formally validated. The objective of the present research was to develop and validate a menu healthiness assessment tool suitable for assessing SMEs in the UK OHFS. Our focus for validation was to assess the tool’s content validity, reliability and predictive validity. As part of the validation process, we applied the tool to sampled SME OHFS outlets in the UK with vs. without a healthier catering accreditation scheme for OHFS businesses.

## 2. Methods

### 2.1. OHFS menu healthiness assessment tool (Menu-Health) development

Database searches (Scopus, Web of Science, Cochrane, Google Scholar) for existing OHFS menu assessment tools were conducted and a snowballing approach was used, by examining citations of relevant papers [38, 42–45, 47, 48] during October-November 2024.

Three published relevant assessment tools / methods for use in the OHFS were identified [38, 42, 45] and criteria from these tools were captured. We supplemented these criteria with current UK government nutritional recommendations for the OHFS in the documents: ‘Healthier Catering Guidance for Different Types of Businesses’ [49], ‘Government Buying Standard for food and catering services’ [50], ‘Calorie reduction. Technical report: guidelines for industry, 2017 baseline calorie levels and the next steps’ [16], ‘Recipe for health: a plan to fix our broken food system’ [51], dietary recommendations in the UK Eatwell Guide [52], and criteria from the Lancashire County Council (LCC) Recipe 4 Health accreditation scheme [53]. LCC are one of 21 County Councils in England with responsibility for public health strategy. The Recipe 4 Health accreditation scheme has been in operation in Lancashire since 2008 and was relaunched in 2019 as part of a childhood obesity prevention programme, and is described in more detail in section 2.5 [54].

A total of 173 health promoting criteria were captured from the existing menu assessment tools and UK government OHFS guidance, and these criteria were grouped into themes (Supplementary Material 1). Criteria were selected for initial inclusion in the menu healthiness assessment tool if they were included in at least one existing tool and/or one guidance document and did not duplicate or align too closely to another criterion. This resulted in 28 unique included criteria relating to six facets of menu healthiness: healthier meal components, healthier drinks, healthier catering practices, portion size, special offers and promotions, diet and nutrition information. We judged each criterion based on whether it could likely be assessed using menus alone (n=20), such as ‘deep fried chips are not available’, or would likely require supplementary data reported by businesses (n=8), such as ‘unsaturated fats are used for frying’.

Alongside each tool criterion, a scoring description was included and examples of evidence required for the criterion to be achieved. The scoring descriptions also signpost resources that may need to be referred to/utilised to assess the criteria, including the UK Eatwell guide [52], the McCance and Widdowson ‘composition of foods integrated dataset’ [55] and the Nutrient Profile Model (NPM) scoring system [56].

### 2.2. Tool scoring

There are four scoring options for each criterion: 1, 0, N/A, and ‘unable to assess’. For each criterion, 1 point is scored if a business meets that criterion. 0 points are scored if a business does not meet that criterion. N/A is scored if the criterion is not applicable to that business. ‘Unable to assess’ is scored if there is insufficient data available to assess that criterion. To enable calculation of a comparable score for individual businesses, we developed a total menu healthiness score as a percentage, with the number of points scored divided by the number of criteria (highest number = 28) assessed *100 (not including any criteria marked N/A or ‘unable to assess’). Higher percentage scores indicated healthier menus.

This approach means that if a business visit and/or menu are the only data source available, the tool can be used to undertake a menu healthiness assessment and by scoring missing criterion (those requiring additional business reporting) using the ‘unable to assess’ option. However, collection of data for all 28 criterion is preferable.

Proposed healthiness scoring categories were 0-49%=poor, 50-70%=fair, ≥71%=good, so that businesses achieving ≥50% of the healthiness criteria were classified as fair or good, and reflecting previous research indicating the majority of food sold in the OHFS would be classified as nutritionally fair or poor [5, 12, 57, 58].

### 2.3. Tool piloting and results

Menu-Health was initially piloted by two of the authors (JB^1^, AF). The researchers independently applied the tool to 10 menus from UK SMEs located in Lancashire, five of which held the LCC Recipe 4 Health award. Adjustments were made to scoring descriptions in the tool and the same 10 menus were re-assessed, resulting in inter-rater agreement levels of 89.29% and a Cohen’s kappa value of 0.85 (strong) [59].

### 2.4. Content validity

Content validity assessment is considered fundamental in instrument development. Content validity was undertaken in three stages: tool development, judgement and quantification, and tool revision and reconstruction [60]. Six researchers and public health practitioners from the UK and Australia were identified for the validation panel on the basis of having developed a similar published measurement tool or having used a similar measurement tool in published research. The validation panel were asked to rate the degree of relevance and clarity of each of the 28 criteria using a five-point scale. For relevance, the five-point scale included the options ‘not relevant’, ‘somewhat relevant’, ‘quite relevant’, ‘relevant’ or ‘highly relevant’. For clarity, the five-point scale included the options ‘not clear’, ‘somewhat clear (needs some modification)’, ‘quite clear (needs minor modification)’, ‘clear’, or ‘very clear’. Following this, validators were also asked to score two statements: ‘The assessment tool provides a comprehensive assessment of the healthiness of the food and drink offering and catering practices of a small-to-medium sized out of home food outlet’ and ‘The assessment tool would be straightforward to use by a trained researcher’ on scales from 0-10, with a score of 0 representing ‘not at all’ and a score of 10 representing ‘extremely’. The validation panel were also asked if any addition criteria should be added to the tool, and for any general comments about the tool.

Validator scores can be found in Supplementary Material 2. Item-level content validity index (I-CVI) was calculated for each criterion, whereby the number of validators rating a criterion as relevant (by selecting ‘quite relevant’, ‘relevant’ or ‘very relevant’) was divided by the total number of validators. Previous research suggests that if a validation panel consists of 5-10 experts, I-CVI ≥0.78 means the criterion should be accepted, and I-CVI <0.78 means the criterion should be revised [60]. Five out of the 28 criteria scored <0.78 and were considered for revision, with amendments agreed between the research team. Three criteria (in the special offers and promotions category) were combined as these were considered to overlap too closely. One criterion (100% fruit juice and/or milk is available) was deleted as it was considered too similar to other criteria, and one further criterion was deleted (staff are trained to suggest and promote healthy/healthier options with less salt, sugar and fat, and higher fibre) as accurate collection of this data was deemed unfeasible. We retained one criterion (relating to availability of no/low alcohol options) as the reasoning provided by the individual validator rating the criterion contradicted UK public health guidance [61], but we altered scoring instructions for the criterion in response to validator feedback. Other adjustments were made to the tool following validator feedback on the clarity of the criteria and scoring system wording, including adding further examples. Post validation, the tool included 26 criteria within six categories, and captured information on food and drink sold, ingredients used, cooking methods, promotions and nutritional information provided. Twenty criteria were assessable by menu analysis alone in most cases, and six criteria were assessable by requesting further information through contact with the business. Tool criteria are shown in Table 1, and the full tool including scoring instructions can be found in Supplementary Material 3.

**Table 1.** Criteria included in final menu healthiness assessment tool

| Criterion number | Category | Criterion |
| --- | --- | --- |
| 1 | Healthier meal components | Non HFSS protein sources (meat/fish/beans/alternative proteins) are available that are not deep fried and are prepared without large amounts of fat or oil |
| 2 | Healthier meal components | Carbohydrate sources are available that are not deep fried and are prepared without large amounts of fat or oil |
| 3 | Healthier meal components | Lower-fat dairy/alternatives are available |
| 4 | Healthier meal components | Grains/wholegrains/fibre-rich carbohydrates are available |
| 5 | Healthier meal components | Vegetable predominant salads are available / meals are offered with side salad |
| 6 | Healthier meal components | Deep-fried chips are not available |
| 7 | Healthier meal components | Vegetable based dishes are offered and/or vegetables are available with meals |
| 8 | Healthier meal components | Fresh fruit is available |
| 9 | Healthier meal components | Children's options are available that contain non-HFSS foods and are prepared without large amounts of fat or oil |
| 10 | Healthier meal components | Desserts are not sold, or if desserts are sold, they are available in smaller sized portions alongside standard sized portions |
| 11 | Healthier drinks | Sugar-sweetened beverages (SSBs) make up no more than 10% of cold beverages offered |
| 12 | Healthier drinks | Water is available |
| 13 | Healthier drinks | No alcoholic drinks are available, or if alcohol is offered, alcohol free and low alcohol equivalents are available |
| 14 | Healthier drinks | Lower-fat dairy/alternatives are offered as standard in hot drinks |
| 15 | Healthier catering practices | Salt offering is minimised |
| 16 | Healthier catering practices | Salt is not added to vegetables, pasta, rice and noodles. Minimal salt is added to potatoes, bread and batters |
| 17 | Healthier catering practices | The default is low/reduced fat spread (unsaturated vegetable oil-based spread or lower fat mayonnaise) OR a small portion / thin scrape of butter / spread OR no spread |
| 18 | Healthier catering practices | Unsaturated fats are used for frying |
| 19 | Healthier catering practices | Items are not fried more than once |
| 20 | Portion size | Meals / items are not available in larger portions (compared to standard/regular size) |
| 21 | Portion size | Meals / items are available in smaller portions (compared to standard/regular size and large) |
| 22 | Special offers and promotions | Special offers and promotions do not include HFSS foods, drinks or options (as defined by NPM score) or items prepared with large amounts of fat or oil |
| 23 | Special offers and promotions | Healthy/healthier options are promoted on menus |
| 24 | Special offers and promotions | Labelled healthy/healthier options are cheaper than less healthy options |
| 25 | Diet and nutrition information | Healthier options are available that cater for different dietary requirements (e.g. vegetarian or vegan or gluten free) |
| 26 | Diet and nutrition information | Calorie information is available on menus |

HFSS= High fat, salt and sugar

To calculate the total tool (scale) content validity index (S-CVI), average I-CVI was used, whereby the number of criteria considered relevant (‘quite relevant’, ‘relevant’ or ‘very relevant’) was divided by the total number of criteria. For S-CVI an Ave-CVI score of ≥0.90 is considered excellent, and 0.80-0.89 acceptable [60, 62]. The tool scored 0.84 for S-CVI. Mean validator scores for comprehensiveness of the tool and ease of use were 8.2 and 5 respectively (out of 10).

### 2.5. Data collection for reliability and predictive validity

To assess tool reliability and predictive validity, we sampled menus and catering practices of 66 OHFS SMEs. To test the predictive validity of the tool, we applied the tool to businesses that held a healthier catering award and similar businesses without the award, hypothesising that businesses with the award would have healthier food, drink and catering practices than those without the award. We also hypothesised that the healthiness of food, drink and catering practices would be associated with business type and IMD decile. It was hypothesised that takeaways would have lower healthiness scores reflecting the findings of previous research [9, 10, 63], and the higher concentration of unhealthy food outlets in more deprived areas [4, 64–66] would result in lower healthiness scores in more deprived areas. We therefore matched businesses with and without the award by business type and IMD decile. The award selected was the local authority healthier catering accreditation scheme ‘Recipe 4 Health’, run by LCC [53].

Recipe 4 Health aims to reward and promote OHFS businesses in the county of Lancashire, North West England, that focus on ‘healthy eating, environmental issues and social responsibility’ [53]. The scheme is available to businesses in the OHFS (cafés, takeaways, restaurants, pubs) and other out of home food providers, including care facilities, prisons, schools and nurseries. The Recipe 4 Health scheme has three tiers: bronze, silver and gold. Businesses must achieve bronze level before progressing to silver, and silver level before progressing to gold. Businesses are eligible to apply for bronze level if they have achieved ≥3 stars in their most recent Food Standards Agency food hygiene inspection [67], silver level if they have achieved ≥4 stars, and gold level if they have achieved 5 stars. Recipe 4 Health has 12 categories: (1) clean (the food hygiene inspection rating), (2) promotions and marketing (of healthier options) (3) fair (foods, prices, weights and measures are described accurately), (4) healthy eating, (5) fruit, fibre and vegetables, (6) salt, (7) sugar, (8) fats and frying, (9) allergens, (10) alcohol awareness, (11) environment (e.g. how waste is disposed of, how much is recycled, how water and electricity are used efficiently) and (12) general (procedures for new staff). The 12 categories encompass 76 essential and 29 desirable criteria across the three tiers. Bronze and silver levels include a mix of essential and desirable criteria; at gold level all criteria are essential.

Current Recipe 4 Health award holders were identified from records shared by the scheme provider. Award holders were matched with similar businesses in Lancashire by business type (café, community café, café in leisure facility, café in children’s play centre, takeaway, pub, restaurant, dessert outlet), cuisine type (mixed, chicken, fish and chip, sandwich, Greek/Turkish/Lebanese, Asian, street food, dessert) and IMD score of business location. Award holders were matched with a business in the same or within two IMD deciles. Food hygiene inspection data was also used when matching businesses to ensure matches met the appropriate eligibility criteria: bronze matches had a food hygiene rating ≥3, silver matches a food hygiene rating ≥4, and gold matches a food hygiene rating of 5. Matched businesses had never applied for the Recipe 4 Health award.

Menus were searched for online and considered eligible for use if posted by the business within the three-month period prior to search date. In the absence of an eligible menu, a researcher visited the business to collect a menu or photographic evidence of the menu. If an eligible online menu could be accessed, the business was contacted to verify the menu was up-to-date, and to collect additional business reported data. This was attempted via phone call (two separate attempts) and if phone contact was unsuccessful, a visit was conducted. Data collection took place from March–June 2025.

### 2.6. Procedure for tool reliability assessment

Two researchers at the University of Liverpool (JB^1^ and a second researcher not involved in tool development) independently applied Menu-Health to 66 small-medium sized OHFS businesses in Lancashire, England. The sample comprised all Recipe 4 Health award holders classed as OHFS businesses (n=33) and matched non-award holders (n=33). The sample comprised 30 cafés, 24 takeaways, 4 restaurants, 4 pubs and 4 dessert outlets. The sample included 11 businesses holding a gold award, 2 businesses holding a silver award and 20 businesses holding a bronze award.

### 2.7. Inter-rater agreement and inter-rater reliability

Inter-rater agreement was calculated as percent agreement, Cohen’s kappa (agreement between the two raters) and Fleiss kappa (agreement between the two raters and the final agreed score), as kappa accounts for chance agreement and is appropriate for use with categorical data [59, 68–70]. There were four categorical variables (1, 0, N/A, unable to assess) and 26 criteria variables. A Cohen’s kappa (κ) of 0-0.20 was considered no agreement, 0.21-0.39 was considered minimal agreement, 0.40-0.59 weak agreement, 0.60-0.79 moderate, 0.80-0.90 strong and >0.90 almost perfect [59]. A Fleiss kappa of <0 was considered poor agreement or no agreement, 0-0.20 slight agreement, 0.21-0.40 fair agreement, 0.41-0.60 moderate agreement, 0.61-0.80 substantial agreement, 0.81-1.00 almost perfect agreement [71].

Intraclass correlation coefficient (ICC) was used to calculate inter-rater reliability, using the healthiness percentage scores for each business from both raters. The 95% confidence interval estimate of ICC was used, with values of < 0.50 considered poor reliability, 0.50-0.75 moderate, 0.75-0.90 good and >0.90 excellent [72].

### 2.8. Predictive validity

Following independent assessments for use in inter-rater agreement analyses, any differences in researcher scores were discussed and an agreed score was reached between the two researchers, by deciding a final score for each criterion for each business for use in predictive validity analyses. Multiple linear regression analyses were performed to evaluate if Recipe 4 Health award status with business type (café/restaurant/pub vs. takeaway/dessert outlet) and IMD decile grouping (IMD1-2 vs. IMD 3-10) was a predictor of menu healthiness. Additional analyses were conducted to examine if award level (silver/gold vs. bronze) predicted menu healthiness.

All analyses were performed using the Psych package in R Statistical Software [73].

## 3. Results

### 3.1. Sample characteristics

Sample characteristics, including business type, cuisine type, IMD decile and award status are shown in Table 2. Takeaway businesses were classified as ‘mixed’ unless one cuisine type was predominantly available (e.g. fish and chips), and more than one of the following were sold: burger, pizza, kebab, curry, fried chicken. Cafés and restaurants were classified as ‘mixed’ unless most dishes originated from a single country or specific cuisine.

**Table 2.** Sample characteristics by business type, cuisine type, IMD decile and award status

|  | Award status |  |  |  |  |  |  |
| --- | --- | --- | --- | --- | --- | --- | --- |
|  | bronze<br>(n) | gold<br>(n) | silver<br>(n) | bronze<br>match<br>(n) | gold<br>match<br>(n) | silver<br>match<br>(n) | Total (n) |
| **Business type** |  |  |  |  |  |  |  |
| Café | 9 | 5 | 1 | 9 | 5 | 1 | 30 |
| Takeaway | 9 | 2 | 1 | 9 | 2 | 1 | 24 |
| Dessert outlet | 1 | 1 | 0 | 1 | 1 | 0 | 4 |
| Restaurant | 1 | 1 | 0 | 1 | 1 | 0 | 4 |
| Pub | 0 | 2 | 0 | 0 | 2 | 0 | 4 |
| Total | 20 | 11 | 2 | 20 | 11 | 2 | 66 |
| **Cuisine type** |  |  |  |  |  |  |  |
| Mixed | 14 | 7 | 1 | 14 | 7 | 1 | 44 |
| Sandwich | 2 | 1 | 0 | 2 | 1 | 0 | 6 |
| Dessert | 1 | 1 | 0 | 1 | 1 | 0 | 4 |
| Asian | 1 | 0 | 0 | 1 | 0 | 0 | 2 |
| Fish and chip | 1 | 0 | 0 | 1 | 0 | 0 | 2 |
| Street food | 1 | 0 | 0 | 1 | 0 | 0 | 2 |
| Chicken | 0 | 0 | 1 | 0 | 0 | 1 | 2 |
| Turkish/Greek/Lebanese | 0 | 2 | 0 | 0 | 2 | 0 | 4 |
| Total | 20 | 11 | 2 | 20 | 11 | 2 | 66 |
| **IMD decile** |  |  |  |  |  |  |  |
| IMD 1-2 | 13 | 2 | 0 | 10 | 6 | 0 | 31 |
| IMD 3-4 | 2 | 6 | 2 | 4 | 4 | 2 | 20 |
| IMD 5-6 | 1 | 2 | 0 | 1 | 0 | 0 | 4 |
| IMD 7-8 | 3 | 1 | 0 | 4 | 0 | 0 | 8 |
| IMD 9-10 | 1 | 0 | 0 | 1 | 1 | 0 | 3 |
| Total | 20 | 11 | 2 | 20 | 11 | 2 | 66 |

IMD= Indices of Multiple deprivation. IMD 1 represents areas with the most deprivation, IMD 10 represents areas with the least deprivation

### 3.2. Inter-rater agreement and inter-rater reliability for business healthiness assessments

Mean agreed healthiness scores were 59.68% (SD 14.29), mean rater 1 healthiness scores were 59.71% (SD 15.50), mean rater 2 healthiness scores were 60.16% (SD 14.48). Inter-rater agreement for healthiness assessment scores was 92.25%, with a Cohen’s kappa of 0.87, indicating strong levels of agreement [59]. The intraclass correlation coefficient for healthiness assessment scores was 0.91, indicating the tool had excellent inter-rater reliability [72]. Individual rater scores, agreed scores, individual rater healthiness categories and agreed healthiness categories for each business can be found in Supplementary Material 4.

### 3.3. Healthiness category assignment reliability

An inter-rater reliability analysis of the healthiness categories assigned by rater 1 and rater 2 scores resulted in a Cohen’s kappa of 0.69 (moderate agreement) [59]. An inter-rater reliability analysis of the healthiness categories assigned by rater 1, rater 2 and agreed scores resulted in a Fleiss kappa of 0.78 (substantial agreement) [71, 74]. A comparison between healthiness category assigned by rater 1, rater 2 and agreed category is shown in Table 3.

**Table 3.**
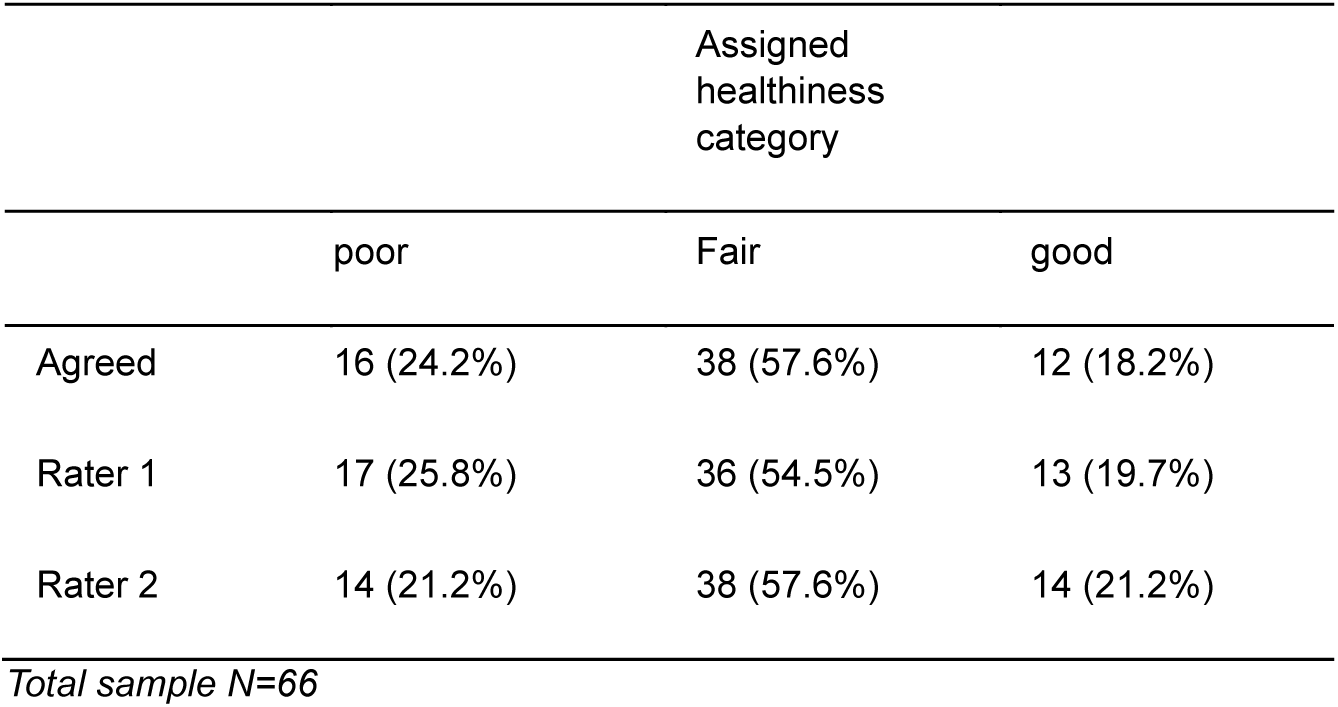
Frequency and percentage of businesses categorised as poor, fair or good

### 3.4. Inter-rater agreement and inter-rater reliability for individual tool criteria and tool categories

Inter-rater percent agreement was calculated for each of the 26 tool criteria. Criterion 21 (meals / items are available in smaller portions compared to standard/regular size and large) and criterion 22 (special offers and promotions do not include HFSS foods, drinks or options, as defined by NPM score, or items prepared with large amounts of fat or oil) had the lowest percent agreement (77.27%). Criterion 2 (carbohydrate sources are available that are not deep fried or do not have large amounts of fat or oil added in the cooking process), criterion 12 (water is available) and criterion 26 (calorie information is available on menus) had the highest percent agreement (100%).

Inter-rater reliability for scores for each of the six tool categories were ĸ=0.87 (healthier meal components), ĸ=0.84 (healthier drinks), ĸ=0.86 (healthier catering practices), ĸ=0.52 (portion size), ĸ=0.46 (special offers and promotions) and ĸ=0.88 (diet and nutrition information).

Analyses were also conducted for each of the 26 tool criteria. Cohen’s kappa values showed that seven criteria had moderate levels of agreement, seven had strong agreement, five had almost perfect agreement and three had perfect agreement. Three criteria (20, 21, 23) had weaker levels of agreement (κ 0.42-0.54) and one criterion (24) had minimal agreement (κ=0.36). The tool categories assessing portion size and special offers and promotions had the lowest percent agreement and inter-rater reliability mean scores. All tool criteria inter-rater percent agreement and reliability values can be found in Supplementary Material 5.

### 3.5. Predictive validity

In 21 of the 33 pairings (63.63%), Recipe 4 Health award holder healthiness scores were numerically higher than matched non-award holders (see Supplementary Material 6). Mean healthiness score for bronze award holders (n=20) was 57.82% (SD 15.76), compared to 55.18% (SD 17.21) for matched businesses (n=20). Mean healthiness score for silver award holders (n=2) was 66.67% (SD 0), compared to 53.81% (SD 8.75) for matched businesses (n=2). Mean healthiness score for gold award holders (n=11) was 69.13% (SD 7.00), compared to 61.62% (SD 8.32) for matched businesses. Mean, maximum and minimum scores for award holders and matched businesses are shown in figure 1.

**Figure 1.**
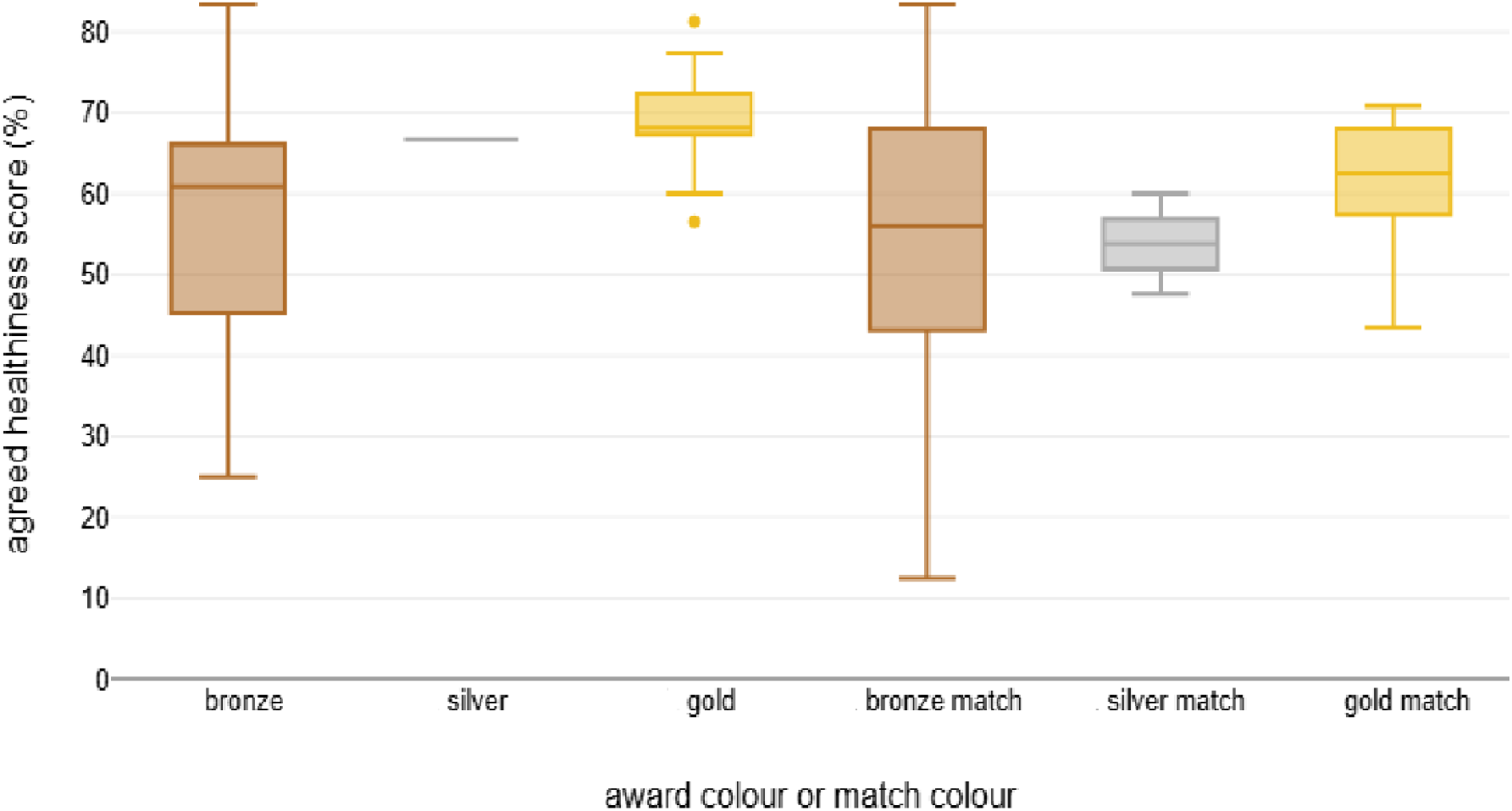
Mean, maximum and minimum healthiness scores of bronze, silver and gold Recipe 4 Health award holders vs. matched non-award holders

### 3.6. Influence of award status, award level, business type and IMD decile on healthiness score

A multiple linear regression was conducted to examine the influence of the predictors award status (award holder or non-award holder), business type and IMD decile on healthiness score (Table 4). Business types were split into two groups for analysis due to imbalance in the sample: cafés, restaurants and pubs were grouped together, and takeaways and dessert outlets were grouped together. IMD deciles were split into two groups for analysis due to imbalance in the sample: IMD 1-2 were grouped together and IMD 3-10 were grouped together. The regression model showed that award status, business type and IMD decile accounted for 29.89% of the variance in the healthiness scores (*F*=8.81, *p*<.001, *R^2^*=0.3). Café/pub/restaurant vs. takeaway/dessert outlet and lower level of deprivation (IMD 3-10) were significant predictors of higher healthiness scores. Award holder status was directionally associated with healthiness score (5% increase in score), but this was not statistically significant. In a model unadjusted for IMD and outlet type, award status results remained the same.

**Table 4.** Impact of award status / award level, business type and IMD decile in predicting healthiness scores

| Predictors | Award status: award holder vs. non-award holder |  |  |  |  |  |  |  |  |  |
| --- | --- | --- | --- | --- | --- | --- | --- | --- | --- | --- |
|  | Model 1* |  |  |  |  | Model 2 |  |  |  |  |
|  | Unstandardised Coefficients (B) | 95% CI | Standard error | t | p | Unstandardised Coefficients (B) | 95% CI | Standard error | t | P |
| Award holder | 4.88 | -1.15, 10.91 | 3.02 | 1.62 | .11 | 4.88 | -2.09, 11.86 | 3.49 | 1.4 | .167 |
| Business type: café/pub/restaurant | 9.45 | 3.21, 15.69 | 3.12 | 3.03 | .004 |  |  |  |  |  |
| IMD decile 3-10 | 9.58 | 3.41, 15.75 | 3.09 | 3.1 | .003 |  |  |  |  |  |
| Predictors | Award level: bronze vs. silver/gold |  |  |  |  |  |  |  |  |  |
|  | Model 1** |  |  |  |  | Model 2 |  |  |  |  |
|  | Unstandardised Coefficients (B) | 95% CI | Standard error | t | p | Unstandardised Coefficients (B) | 95% CI | Standard error | t | P |
| Bronze level | 3.09 | -3.99, 10.18 | 3.54 | 0.87 | .386 | 0.58 | -7.22, 8.37 | 3.9 | 0.15 | .883 |
| Silver/gold level | 7.63 | -0.68, 15.95 | 4.16 | 1.84 | .071 | 11.5 | 2.5, 20.51 | 4.51 | 2.55 | .013 |
| Business type: café/pub/restaurant | 9.27 | 3.01, 15.52 | 3.13 | 2.96 | .004 |  |  |  |  |  |
| IMD decile 3-10 | 8.43 | 1.81, 15.05 | 3.31 | 2.55 | .013 |  |  |  |  |  |

Model 1 examined the impact of the predictors award status\* or award level\*\*, business type and IMD decile on healthiness score. Model 2 was unadjusted for business type or IMD decile

A multiple linear regression was conducted to examine the influence of award level (bronze or silver/gold) on healthiness score (Table 4). Silver and gold levels were grouped together due to the small number of silver award holders. An unadjusted model examined the influence of bronze and silver/gold award level on healthiness score. This model was statistically significant, accounting for 10.05% of the variance in healthiness score (*F*=3.52, *p*=.035, *R^2^*=0.1). In this model, silver/gold award level vs. no award was a significant predictor of healthiness score, but bronze level award vs. no award was not.

In an adjusted model, award level (bronze or silver/gold), business type and IMD decile accounted for 30.94% of the variance in the healthiness scores, which was a significant effect (*F*=6.83, *p*<.001, *R^2^*=0.31). The individual predictors were further examined and indicated that being a café/pub/restaurant and being located in IMD decile 3-10 were significant predictors of healthiness score, but having the bronze level award or the silver/gold level award were not statistically significant predictors of healthiness score in this model.

Adjusted analysis models were pre-registered on the Open Science Framework https://osf.io/a743g/?view_only=a55924b192004570936f7bd385b1e902. All tests met assumptions for multicollinearity and heteroskedasticity.

### 3.7. Sensitivity analyses

Exploratory analyses were conducted removing the four tool criteria with a Cohen’s kappa value <0.60 (classified as weak and minimal agreement for inter-rater reliability). Mean healthiness scores for all groups increased with the four criteria removed (mean group increase 6.05%). Significance of all regression results remained the same as in the main analyses. Further exploratory analyses were conducted removing the six criteria that required additional contact with the business to undertake assessment. Mean healthiness scores for all groups decreased with the six criteria removed (mean group decrease 6.35%). Significance of regression results remained the same as in the main analyses. Full results from exploratory analyses can be found in Supplementary Material 7. Across all sensitivity analyses, overall inter-rater agreement remained high.

## 4. Discussion

We developed a 26-criteria tool, Menu-Health, to assess the healthiness of menus in SMEs in the UK OHFS based on nutritional recommendations for the UK OHFS. Content validity of the tool was assessed by an expert panel and we conducted formal analyses on tool inter-rater agreement, reliability and predictive validity. Analysis of business healthiness scores indicated Menu-Health had strong levels of inter-rater agreement (92%) and excellent inter-rater reliability (ĸ=0.87, ICC=0.91), suggesting the tool is an accurate and reliable measure of menu healthiness. Consistent with hypotheses, takeaways and dessert outlets and businesses located in areas of greater deprivation predicted lower healthiness scores. These findings align with previous research observing the poor nutritional profile of takeaway food, and the increased availability of less healthy food in more deprived areas [9, 10, 65, 75], and confirm the predictive validity of the tool.

Menu healthiness scores for businesses holding a healthier catering award were on average higher than those without the award (5% higher score), but this difference was not statistically significant. When different award levels were examined, outlets with higher award levels had significantly higher healthiness scores than matched non-award holders (+8% at gold level), but statistical significance depended on adjustment for outlet type and local area deprivation. A possible explanation for the relatively small difference observed between award holders vs. non-award holders is that the type of businesses applying for accreditation awards are more likely to already have healthier food, drink and catering practices, and the scheme does not result in menu changes in order to receive accreditation [21, 24]. Furthermore, it is important to note that the business accreditation scheme examined (Recipe 4 Health) [53], included a range of factors separate to healthiness of catering practices, such as using environmentally sustainable practices, controlling allergens, and accurately labelling food and drink items.

Comparison with other tools is limited as few tools designed specifically to assess the OHFS have formally measured content validity and reliability [76]. A US tool (NEMS-R), had similar inter-rater agreement scores, but inter-rater reliability scores were higher for Menu-Health [38]. Agreement and reliability for Menu-Health were also significantly higher than for one tool designed to assess restaurants in three urban areas of China and compare favourably with a fast-food restaurant assessment observation checklist developed in the US [77].

The two criteria assessing portion size (20 and 21) and three criteria assessing promotion of healthier options (22, 23 and 24) had the lowest inter-rater reliability scores (ĸ 0.36-0.6).

Findings were similar with the NEMS-R tool, where availability of reduced-size portions (ĸ=0.27) and healthy options being highlighted/encouraged (ĸ=0.33) had the lowest inter-rater reliability scores [38]. These similar findings would suggest that objective assessment of portion size and healthier options is more challenging than other criteria. Adjusting the tool to remove the four criteria with the lowest inter-rater reliability scores resulted in marginally higher healthiness assessment scores in all award holder and matched groups (mean group healthiness score increase 6%), but results of predictive validity analyses remained unchanged and levels of overall inter-rater agreement and reliability were largely comparable.

Menu-Health requires contact with businesses to be used in full, as scoring of six of the 26 tool criteria requires information not typically available from menus. Analyses in which we removed the six criteria requiring additional contact with businesses resulted in lower average healthiness assessment scores (-6%), but predictive validity analyses remained unchanged, and levels of overall inter-rater agreement and reliability were largely comparable. This suggests that the tool can likely be used to assess healthiness of menus when contact with businesses is not feasible. However, full assessments with business contact are recommended to avoid businesses receiving inflated healthiness scores due to missing data.

Healthiness scores of assessed outlets ranged from 13% to 83%. Most businesses (58%) were classed as ‘fair’ and more were classed as ‘poor’ (24%) than ‘good’ (18%). These data highlight that healthier catering practices in SMEs in the UK OHFS need improvement. This is important because greater availability and promotion of healthier, more nutritious food and drink is needed to tackle key public health challenges such as rising obesity prevalence and associated health and economic consequences [78, 79].

Limitations of the study include the relatively small number of businesses used to assess the predictive validity of the tool. The sample size was dictated by the number and type of OHFS businesses that held the Recipe 4 Health award and was predominantly comprised of mixed cuisine cafés and takeaways. The majority (45/66) of businesses were located in IMD deciles 1-3, indicating relatively high levels of deprivation. Similar to findings in other research, business assessments were relatively time-consuming to undertake (25-30 minutes per business) [38, 41, 77], and although additional information was requested via phone call in the first instance, a number of business visits were also required to collect additional data.

Validator feedback on the comprehensiveness of Menu-Health was positive (mean score 8/10), however initial ratings for tool ease of use prior to tool finalization were average (mean score 5/10). To address this tool training was used to facilitate accurate and consistent assessment between two researchers, and both researchers were able to use the tool with relative ease. The tool was developed using UK nutritional guidance and recommendations for the UK OHFS and tested in specific SME outlets within one region of the UK, so generalisability to other regions/countries and outlet types would now be valuable, including in larger OHFS outlets that contribute significantly to market share in the sector [20]. It is also unclear at present whether the tool could be used in non-research contexts (e.g., public health practitioners), given that ease of use may be a potential limitation of the tool. Further validation work with other users of the tool is now warranted.

Twenty two of the 26 (85%) tool criteria had moderate, strong, almost perfect or perfect inter-rater reliability. However, three criteria relating to portion size, special offers and promotions (20, 21, 23) had weak agreement for inter-rater reliability (ĸ=0.42-0.54) and one criterion (24) ‘labelled healthy/healthier options are cheaper than less healthy options’, had high levels of inter-rater percent agreement (91%) but minimal agreement for inter-rater reliability (ĸ=0.36). We opted to retain these criteria due to their relevance to assessing menu healthiness.

Therefore, it is important to note that although the overall tool and the tool sub categories produce reliable scoring of menu healthiness, some of the individual tool criteria may benefit from refinement to improve reliability of scoring.

Strengths of the study include the robust assessment of tool content validity, reliability and predictive validity. Strengths of the tool include the comprehensiveness of assessment of OHFS menu healthiness, including the availability of healthier food and drink options for adults and children, ingredients used, cooking methods, portions sizes, special offers, promotions and the availability of nutritional information. Menu-Health can also be used to assess a range of SME OHFS outlets and cuisine types which are historically difficult to assess due to limited validated tools.

## 5. Conclusion

The present research developed a reliable, validated, comprehensive tool for assessing the healthiness of food, drink and catering practices in UK OHFS SMEs. Potential applications of the tool include monitoring the healthiness of catering practices and evaluating interventions designed to improve SME menu healthiness.

## Data Availability

All data produced in the present study are available upon reasonable request to the authors

## Acknowledgements

We would like to thank Francesca Quigley for assisting with tool reliability assessments.

## Conflicts of Interest

All authors declare no conflicts of interest with the publication of this research paper.

## Supplementary Materials

The development and validation of a menu healthiness assessment tool (Menu-Health) for small-to-medium sized businesses in the UK out-of- home food sector

## Exploratory analysis 1: Removing four criteria with lowest inter-rater reliability scores

Mean healthiness scores for all groups increased with the four criteria removed (mean group increase 6.05%): bronze award holders increased by 6.4% (matches by 6.32%), silver award holders by 5.16% (matches by 3.01%) and gold award holders by 7.91% (matches by 7.5%, figure 1).

**Figure S1.**
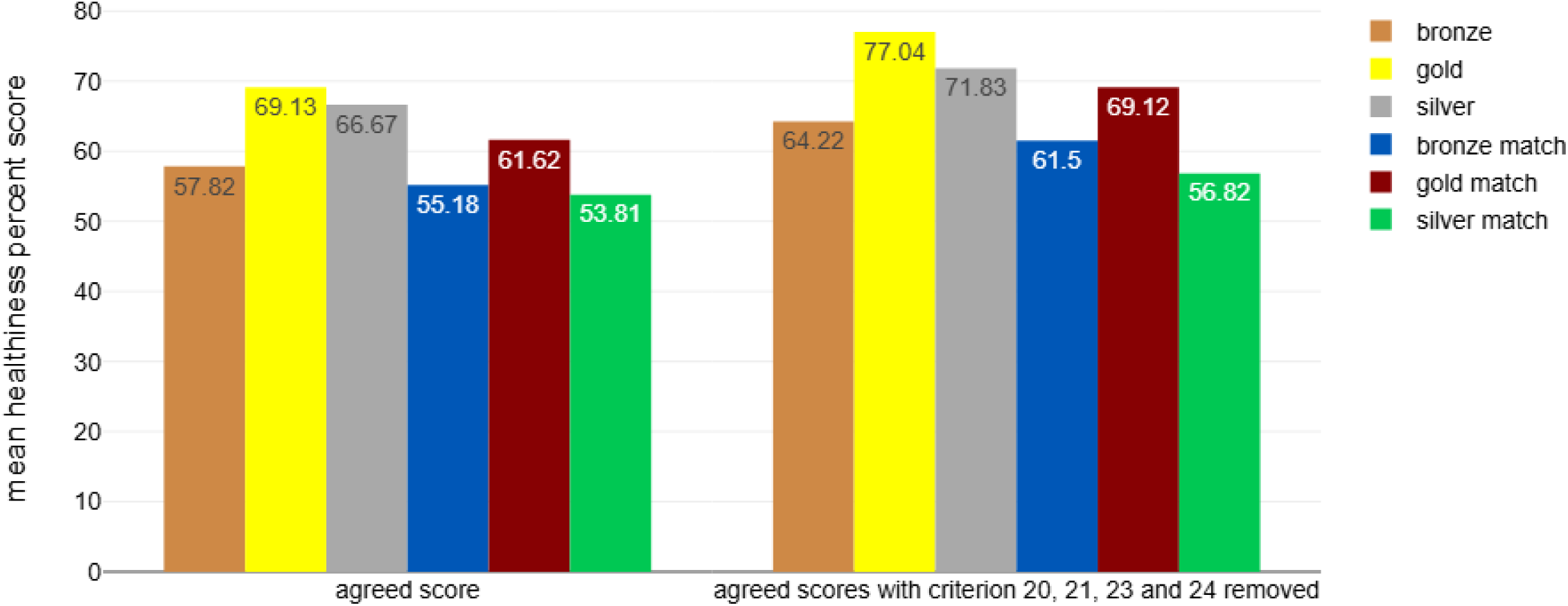
Comparison of group mean healthiness scores when all tool criteria are included (on left) and with four criteria with lowest inter-rater reliability scores removed (on right)

The same regression models were conducted as in the main analyses but with tool criteria 20, 21, 23 and 24 (Cohen’s kappa <0.60) removed. Firstly, a multiple linear regression was conducted to examine the influence of the predictors award status (award holder or non-award holder), business type and IMD decile on healthiness score. The regression model showed the three predictors accounted for 29.75% of the variance in the healthiness scores, which was a significant effect (*F*=8.75, *p* <.001, *R^2^*=0.3). The individual predictors were further examined and indicated that being a café/pub/restaurant (t=3.18, p= .002) and being located in IMD 3-10 (t=2.94, p=.005) were significant predictors of healthiness score in this model, but being a Recipe 4 Health award holder was not (t=1.57, p=.122).

Secondly, a multiple linear regression was conducted to examine the influence of the predictors award level (bronze or silver/gold), business type and IMD decile on healthiness score. The regression model showed the four predictors accounted for 30.9% of the variance in the healthiness scores, which was a significant effect (*F*=6.82, *p*<.001, *R^2^*=0.31). The individual predictors were further examined and indicated that being a café/pub/restaurant (t=3.12, p=.003) and being located in IMD decile 3-10 (t=2.28, p=.02) were significant predictors of healthiness score in this model, but having the bronze level award (t=0.81, p=.424) or the silver/gold level award (t=1.83, p=.072) were not. The significance of exploratory regression analysis results with tool criteria 20, 21, 23 and 24 removed was no different to main regression analysis results.

## Exploratory analysis 2: Removing six criteria that require additional contact with businesses to undertake assessment

Mean healthiness scores for all groups decreased with the six criteria removed (mean group decrease 6.35%): bronze award holders decreased by 6.49% (matches by 4.74%), silver award holders by 7.99% (matches by 4.27%) and gold award holders by 7.35% (matches by 7.34%, figure 2).

**Figure S2.**
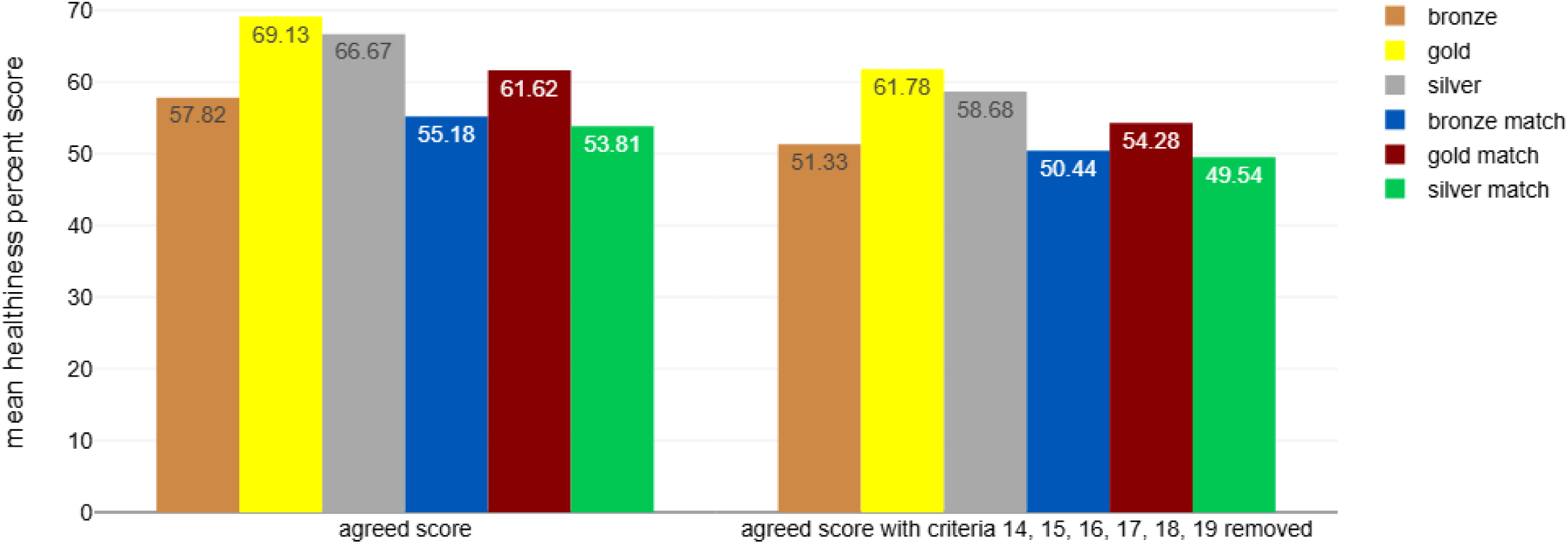
Comparison of group mean healthiness scores when all tool criteria are included (on left) and with six criteria requiring additional contact with business removed (on right)

The same regression models were conducted as in the main analyses but with the tool criteria that required additional contact with businesses removed (criteria 14, 15, 16, 17, 18 and 19). Firstly, a multiple linear regression was conducted to examine the influence of the predictors award status (award holder or non-award holder), business type and IMD decile on healthiness score. The regression model showed the three predictors accounted for 33.09% of the variance in the healthiness scores, which was a significant effect (*F*=10.22, *p* <.001, *R^2^*=0.33). The individual predictors were further examined and indicated that being a café/pub/restaurant (t=3.51, p= .001) and being located in IMD 3-10 (t=3.28, p=.002) were significant predictors of healthiness score in this model, but being a Recipe 4 Health award holder was not (t=1.21, p=.232).

Secondly, a multiple linear regression was conducted to examine the influence of the predictors award level (bronze or silver/gold), business type and IMD decile on healthiness score. The regression model showed the four predictors accounted for 33.5% of the variance in the healthiness scores, which was a significant effect (*F*=7.68, *p*<.001, *R^2^*=0.33). The individual predictors were further examined and indicated that being a café/pub/restaurant (t=3.45, p=.001) and being located in IMD decile 3-10 (t=2.82, p=.006) were significant predictors of healthiness score in this model, but having the bronze level award (t=0.7, p=.486) or the silver/gold level award (t=1.29, p=.201) were not. The significance of exploratory regression analysis results with tool criteria 14-19 removed was no different to main regression analysis results.

---

**Supplementary Material 1.** Captured criteria from existing tools / UK recommendations for OHFS

| Item description | Method of assessment | Methodology | Category | Applicable to which outlets? | Relevance to SME? | Author / Source |
| --- | --- | --- | --- | --- | --- | --- |
| Number of special offers, meal deals, sharing meals, set meals | Menu analysis | Count number | Special offers | Takeaway / Restaurant / Café | Y | Goffe (2020) |
| Number of meals available in multiple sizes | Menu analysis | Count number | Multiple sizes | Takeaway / Restaurant / Café | Y | Goffe (2020) |
| Number of meal items labelled as small | Menu analysis | Count number | Smaller portions | Takeaway / Restaurant / Café | Y | Goffe (2020) |
| Number of items with health related labels | Menu analysis | Count number | Healthy / healthier options | Takeaway / Restaurant / Café | Y | Goffe (2020) |
| Number of dessert items | Menu analysis | Count number | Desserts | Takeaway / Restaurant / Café | Y | Goffe (2020) |
| Count of all mentions of salad or related items (e.g. coleslaw) | Menu analysis | Count number | Salads | Takeaway / Restaurant / Café | Y | Goffe (2020) |
| Count of all mentions of chips / fries / wedges | Menu analysis | Count number | Chips | Takeaway / Restaurant / Café | Y | Goffe (2020) |
| Number of different vegetables mentioned | Menu analysis | Count number | Vegetables | Takeaway / Restaurant / Café | Y | Goffe (2020) |
| Number of different dietary requirements explicitly catered for | Menu analysis | Count number | Dietary requirements | Takeaway / Restaurant / Café | Y | Goffe (2020) |
| Number of nutritional metrics e.g. kcal | Menu analysis | Count number | Nutritional information | Takeaway / Restaurant / Café | Y (not mandatory in independent OHFS) | Goffe (2020) |
| Number of sugar-sweetened beverages | Menu analysis | Count number | Sugar-sweetened beverages | Takeaway / Restaurant / Café | Y | Goffe (2020) |
| Number of diet drinks | Menu analysis | Count number | Diet drinks | Takeaway / Restaurant / Café | Y | Goffe (2020) |
| Number of milk (non-sweetened) options | Menu analysis | Count number | Dairy/alternatives | Takeaway / Restaurant / Café | Y | Goffe (2020) |
| Number of water options | Menu analysis | Count number | Water | Takeaway / Restaurant / Café | Y | Goffe (2020) |
| Number of alcoholic drinks | Menu analysis | Count number | Alcohol | Takeaway / Restaurant / Café | Y (not all businesses) | Goffe (2020) |
| Presence of vegetable based mixed dishes | Menu analysis | Compare number of healthy vs. unhealthy | Vegetables | Takeaway / Restaurant / Café | Y | Pulker (MAST, 2023) |
| Presence of fruit | Menu analysis | Compare number of healthy vs. unhealthy | Fruit | Takeaway / Restaurant / Café | Y | Pulker (MAST, 2023) |
| Presence of grains / cereals | Menu analysis | Compare number of healthy vs. unhealthy | Grains / fibre | Takeaway / Restaurant / Café | Y | Pulker (MAST, 2023) |
| Presence of meat / alternatives | Menu analysis | Compare number of healthy vs. unhealthy | Meat/alternatives | Takeaway / Restaurant / Café | Y | Pulker (MAST, 2023) |
| Presence of dairy / alternatives | Menu analysis | Compare number of healthy vs. unhealthy | Dairy/alternatives | Takeaway / Restaurant / Café | Y | Pulker (MAST, 2023) |
| Presence of SSBs / energy drinks / confectionary | Menu analysis | Compare number of healthy vs. unhealthy | Sugar-sweetened beverages | Takeaway / Restaurant / Café | Y | Pulker (MAST, 2023) |
| Availability of health main dishes / starters | Menu analysis | Y/N? | Healthy / healthier options | Takeaway / Restaurant / Café | Y | Saelens (NEMS-R, 2007) |
| Healthy options identified on menus | Menu analysis | Y/N? | Healthy / healthier options | Takeaway / Restaurant / Café | Y | Saelens (NEMS-R, 2007) |
| Availability of salads as main dishes | Menu analysis | Y/N? | Salads | Takeaway / Restaurant / Café | Y | Saelens (NEMS-R, 2007) |
| Availability of main dish salad options considered healthy | Menu analysis | Y/N? | Healthy / healthier options | Takeaway / Restaurant / Café | Y | Saelens (NEMS-R, 2007) |
| Availability of fruit | Menu analysis | Y/N? | Fruit | Takeaway / Restaurant / Café | Y | Saelens (NEMS-R, 2007) |
| Availability of non-fried vegetables | Menu analysis | Y/N? | Vegetables | Takeaway / Restaurant / Café | Y | Saelens (NEMS-R, 2007) |
| Availability of baked chips | Menu analysis | Y/N? | Chips | Takeaway / Restaurant / Café | Y | Saelens (NEMS-R, 2007) |
| Availability of whole grain bread | Menu analysis | Y/N? | Grains / fibre | Takeaway / Restaurant / Café | Y | Saelens (NEMS-R, 2007) |
| Availability of diet soda | Menu analysis | Y/N? | Diet drinks | Takeaway / Restaurant / Café | Y | Saelens (NEMS-R, 2007) |
| Availability of 100% fruit juice | Menu analysis | Y/N? | Fruit juice | Takeaway / Restaurant / Café | Y | Saelens (NEMS-R, 2007) |
| Availability of 1% or non-fat milk | Menu analysis | Y/N? | Dairy/alternatives | Takeaway / Restaurant / Café | Y | Saelens (NEMS-R, 2007) |
| Nutrition information on menu | Menu analysis | Y/N? | Nutritional information | Takeaway / Restaurant / Café | Y (not mandatory in independent OHFS) | Saelens (NEMS-R, 2007) |
| Availability of reduced-size portions | Menu analysis | Y/N? | Smaller portions | Takeaway / Restaurant / Café | Y | Saelens (NEMS-R, 2007) |
| Special requests encouraged / discouraged | Menu analysis / site visit / questionnaire | Y/N? | Special requests | Takeaway / Restaurant / Café | Y | Saelens (NEMS-R, 2007) |
| Presence of salad bar | Menu analysis / site visit / questionnaire | Y/N? | Salads | Takeaway / Restaurant / Café | Y | Saelens (NEMS-R, 2007) |
| Large portions encouraged (on menu) | Menu analysis | Y/N? | Large portions | Takeaway / Restaurant / Café | Y | Saelens (NEMS-R, 2007) |
| Overeating encouraged (on menu) | Menu analysis | Y/N? | Overconsumption | Takeaway / Restaurant / Café | Y | Saelens (NEMS-R, 2007) |
| Presence of 'all you can eat' or 'unlimited' options | Menu analysis | Y/N? | Overconsumption | Takeaway / Restaurant / Café | Y | Saelens (NEMS-R, 2007) |
| Promotion of low-carbohydrate options | Menu analysis | Y/N? | Healthy / healthier options | Takeaway / Restaurant / Café | Y | Saelens (NEMS-R, 2007) |
| Pricing: combination meal cheaper than sum price of individual items | Menu analysis | Y/N? | Price promotions | Takeaway / Restaurant / Café | Y | Saelens (NEMS-R, 2007) |
| Pricing: healthy starter cheaper than regular | Menu analysis | Y/N? | Price promotions | Takeaway / Restaurant / Café | Y | Saelens (NEMS-R, 2007) |
| No charge for shared starter | Menu analysis | Y/N? | Price promotions | Takeaway / Restaurant / Café | Y | Saelens (NEMS-R, 2007) |
| Pricing: small portion cheaper than regular portion | Menu analysis | Y/N? | Price promotions | Takeaway / Restaurant / Café | Y | Saelens (NEMS-R, 2007) |
| Presence of nutrition information near point-of-purchase | site visit / questionnaire | Y/N? | Nutritional information | Takeaway / Restaurant / Café | Y (not mandatory in independent OHFS) | Saelens (NEMS-R, 2007) |
| Signs highlighting healthy options | site visit / questionnaire | Y/N? | Signage (healthy) | Takeaway / Restaurant / Café | Y | Saelens (NEMS-R, 2007) |
| Signs encouraging overeating | site visit / questionnaire | Y/N? | Signage (unhealthy) | Takeaway / Restaurant / Café | Y | Saelens (NEMS-R, 2007) |
| Signs encouraging healthy eating | site visit / questionnaire | Y/N? | Signage (healthy) | Takeaway / Restaurant / Café | Y | Saelens (NEMS-R, 2007) |
| Signs encouraging unhealthy eating | site visit / questionnaire | Y/N? | Signage (unhealthy) | Takeaway / Restaurant / Café | Y | Saelens (NEMS-R, 2007) |
| If child's meal available, is not fried option and served with milk/fruit juice | Menu analysis | Y/N? | Children | Takeaway / Restaurant / Café | Y | Saelens (NEMS-R, 2007) |
| Make small / half portions available and market to everyone | Menu analysis / site visit / questionnaire | Y/N? | Smaller portions | Takeaway / Restaurant / Café | Y | Healthier Catering Guidance for Different Types of Businesses (PHE, 2019) |
| Offer reduced-size specials | Menu analysis / site visit / questionnaire | Y/N? | Smaller portions | Takeaway / Restaurant / Café | Y | Healthier Catering Guidance for Different Types of Businesses (PHE, 2019) |
| Use plain, uncoated, medium or thick (at least 14mm), straight cut chips | site visit / questionnaire | Y/N? | Chips | Fish & Chip shop / Deep friers | Y | Healthier Catering Guidance for Different Types of Businesses (PHE, 2019) |
| Fry at 160-165°C (high efficiency fryer) or below 175°C (traditional fryer) | site visit / questionnaire | Y/N? | fats and frying | Fish & Chip shop / Deep friers | Y | Healthier Catering Guidance for Different Types of Businesses (PHE, 2019) |
| Bang, shake and drain oil | site visit / questionnaire | Y/N? | fats and frying | Fish & Chip shop / Deep friers | Y | Healthier Catering Guidance for Different Types of Businesses (PHE, 2019) |
| Fry with a liquid oil (sunflower / rapeseed) | site visit / questionnaire | Y/N? | fats and frying | Fish & Chip shop / Deep friers | Y | Healthier Catering Guidance for Different |
|  |  |  |  |  |  | Types of Businesses (PHE, 2019) |
| Only add salt if customers request it | site visit / questionnaire | Y/N? | Salt | Fish & Chip shop / Deep friers | Y | Healthier Catering Guidance for Different Types of Businesses (PHE, 2019) |
| Use a salt shaker with fewer holes (max 5 holes) | site visit / questionnaire | Y/N? | Salt | Fish & Chip shop / Deep friers | Y | Healthier Catering Guidance for Different Types of Businesses (PHE, 2019) |
| Do not add salt to batter mix / no added salt in pre-prepared batter mix | site visit / questionnaire | Y/N? | Salt | Fish & Chip shop / Deep friers | Y | Healthier Catering Guidance for Different Types of Businesses (PHE, 2019) |
| Healthier drinks are default option (water, lower fat milks, low cal or no added sugar) | Menu analysis / site visit / questionnaire | Y/N? | Drinks | Takeaway / Restaurant / Café | Y | Healthier Catering Guidance for Different Types of Businesses (PHE, 2019) |
| Include healthier drinks / salad / vegetables and a piece of fruit with meal deals | Menu analysis / site visit / questionnaire | Y/N? | Special offers | Takeaway / Restaurant / Café | Y | Healthier Catering Guidance for Different Types of Businesses (PHE, 2019) |
| Use wholegrain/brown/50:50 bread | Menu analysis / site visit / questionnaire | Y/N? | Grains / fibre | Takeaway / Restaurant / Café | Y | Healthier Catering Guidance for Different Types of Businesses (PHE, 2019) |
| Use lower salt bread (less than 0.9mg salt per 100g) | site visit / questionnaire | Y/N? | Salt | Takeaway / Restaurant / Café | Y | Healthier Catering Guidance for Different Types of Businesses (PHE, 2019) |
| Use low / reduced fat spread (ideally from unsaturated vegetable oil) or thin scrape /no spread | site visit / questionnaire | Y/N? | Fats | Takeaway / Restaurant / Café | Y | Healthier Catering Guidance for Different Types of Businesses (PHE, 2019) |
| Use spread with less than 1.06g salt per 100g | site visit / questionnaire | Y/N? | Fats | Takeaway / Restaurant / Café | Y | Healthier Catering Guidance for Different Types of Businesses (PHE, 2019) |
| Use reduced-fat hard cheese / edam / mozzarella / lower fat cream cheese | Menu analysis / site visit / questionnaire | Y/N? | Dairy/alternatives | Takeaway / Restaurant / Café | Y | Healthier Catering Guidance for Different Types of Businesses (PHE, 2019) |
| Use lower fat and salt mayonnaise / dressing and low-fat plain yoghurt | Menu analysis / site visit / questionnaire | Y/N? | fats | Takeaway / Restaurant / Café | Y | Healthier Catering Guidance for Different Types of Businesses (PHE, 2019) |
| Check any pre-made sandwich fillings are lower in salt, sugar, fat and saturated fat | site visit / questionnaire | Y/N? | Healthy / healthier options | Takeaway / Restaurant / Café | Y | Healthier Catering Guidance for Different Types of Businesses (PHE, 2019) |
| Use lean meats without skin and cut off any visible fat | site visit / questionnaire | Y/N? | Meat/alternatives | Takeaway / Restaurant / Café | Y | Healthier Catering Guidance for Different Types of Businesses (PHE, 2019) |
| Use less processed meat and choose those with lower salt and fat content | Menu analysis / site visit / questionnaire | Y/N? | Meat/alternatives | Takeaway / Restaurant / Café | Y | Healthier Catering Guidance for Different Types of Businesses (PHE, 2019) |
| Offer salad in or with every sandwich | Menu analysis / site visit / questionnaire | Y/N? | Salads | Takeaway / Restaurant / Café | Y | Healthier Catering Guidance for Different Types of Businesses (PHE, 2019) |
| Remove confectionary, cakes, pastries, biscuits, morning goods and sugary drinks from till points | site visit / questionnaire | Y/N? | Positional promotions | Takeaway / Restaurant / Café | Y | Healthier Catering Guidance for Different Types of Businesses (PHE, 2019) |
| Offer desserts in smaller portions. Choose products that meet sugar / calorie reduction guidelines | Y/N? | Desserts | Desserts | Takeaway / Restaurant / Café | Y | Healthier Catering Guidance for Different Types of Businesses (PHE, 2019) |
| Promote healthier sandwich options, snacks and drinks with meal deals (e.g. fresh fruit, plain nuts, low-fat yoghurts, baked/popped crisps, smaller packets, small portions of biscuits/cakes/pastries, or ones that meet sugar / calorie reduction guidelines) | site visit / questionnaire | Y/N? | Special offers | Takeaway / Restaurant / Café | Y | Healthier Catering Guidance for Different Types of Businesses (PHE, 2019) |
| Put fruit and healthy snacks in prominent areas / in store / window displays | site visit / questionnaire | Y/N? | Positional promotions | Takeaway / Restaurant / Café | Y | Healthier Catering Guidance for Different Types of Businesses (PHE, 2019) |
| Put healthier drinks at eye-level in the fridge | site visit / questionnaire | Y/N? | Positional promotions | Takeaway / Restaurant / Café | Y | Healthier Catering Guidance for Different Types of Businesses (PHE, 2019) |
| Limit fried foods, offer steamed, boiled or grilled dishes instead | Menu analysis | Y/N? | Fats | Takeaway / Restaurant / Café | Y | Healthier Catering Guidance for Different Types of Businesses (PHE, 2019) |
| Avoid frying food more than once | site visit / questionnaire | Y/N? | fats and frying | Takeaway / Restaurant / Café | Y | Healthier Catering Guidance for Different Types of Businesses (PHE, 2019) |
| Reduce the amount of salt and salty ingredients e.g. salt, MSG, soy sauce, hoi sin sauce, oyster sauce, stock, ketchup gradually over time | site visit / questionnaire | Y/N? | Salt | Takeaway / Restaurant / Café | Y | Healthier Catering Guidance for Different Types of Businesses (PHE, 2019) |
| Remove salt / soy sauce from tables | site visit / questionnaire | Y/N? | Salt | Takeaway / Restaurant / Café | Y | Healthier Catering Guidance for Different Types of Businesses (PHE, 2019) |
| Avoid adding salt to vegetables, potatoes, pasta, rice, noodles, breads and spices during cooking | site visit / questionnaire | Y/N? | Salt | Takeaway / Restaurant / Café | Y | Healthier Catering Guidance for Different Types of Businesses (PHE, 2019) |
| Add vegetables / pulses to as many dishes as possible, including to boiled rice / more vegetables on pizzas | Menu analysis | Y/N? | Vegetables | Takeaway / Restaurant / Café | Y | Healthier Catering Guidance for Different Types of Businesses (PHE, 2019) |
| Offer brown rice / wholemeal chapatis / wholewheat pasta | Menu analysis | Y/N? | Grains / fibre | Takeaway / Restaurant / Café | Y | Healthier Catering Guidance for Different Types of Businesses (PHE, 2019) |
| Promote healthier options in meal deals | Menu analysis | Y/N? | Special offers | Takeaway / Restaurant / Café | Y | Healthier Catering Guidance for Different Types of Businesses (PHE, 2019) |
| Train staff to suggest and promote healthier options with higher fibre, less salt, sugar and fat | site visit / questionnaire | Y/N? | Healthy promotions | Takeaway / Restaurant / Café | Y | Healthier Catering Guidance for Different Types of Businesses (PHE, 2019) |
| Add a 'healthier options' section / page to menu to help customers identify healthy options | Menu analysis | Y/N? | Healthy / healthier options | Takeaway / Restaurant / Café | Y | Healthier Catering Guidance for Different Types of Businesses (PHE, 2019) |
| Do not overfill containers / use slightly smaller containers | site visit / questionnaire | Y/N? | Smaller portions | Takeaway | Y | Healthier Catering Guidance for Different Types of Businesses (PHE, 2019) |
| Cook in the tandoor rather than deep frying, where possible | site visit / questionnaire | Y/N? | Fats | Takeaway | Y | Healthier Catering Guidance for Different Types of Businesses (PHE, 2019) |
| Do not serve poppadoms free of charge | site visit / questionnaire | Y/N? | Price promotions | Takeaway / Restaurant / Café | Y | Healthier Catering Guidance for Different Types of Businesses (PHE, 2019) |
| Offer half sized portions (e.g. of pizza or pasta) with a side salad and a tea/coffee | Menu analysis | Y/N? | Smaller portions | Takeaway / Restaurant / Café | Y | Healthier Catering Guidance for Different |
|  |  |  |  |  |  | Types of Businesses (PHE, 2019) |
| Use less cream and swap to half-fat crème fraîche, single cream or low fat plain yoghurt instead | site visit / questionnaire | Y/N? | Dairy/alternatives | Takeaway / Restaurant / Café | Y | Healthier Catering Guidance for Different Types of Businesses (PHE, 2019) |
| Offer salads with dressings on the side | Menu analysis / site visit / questionnaire | Y/N? | Healthy / healthier options | Takeaway / Restaurant / Café | Y | Healthier Catering Guidance for Different Types of Businesses (PHE, 2019) |
| Use less butter on items such as garlic bread and brush with olive oil instead | site visit / questionnaire | Y/N? | Fats | Takeaway / Restaurant / Café | Y | Healthier Catering Guidance for Different Types of Businesses (PHE, 2019) |
| Make it clear on the menu how many people a pizza will serve, create more slices on a pizza | Menu analysis | Y/N? | Smaller portions | Takeaway / Restaurant / Café | Y | Healthier Catering Guidance for Different Types of Businesses (PHE, 2019) |
| Avoid offering meat or cheese stuffed crust pizzas | Menu analysis | Y/N? | Healthy / healthier options | Takeaway / Restaurant / Café | Y | Healthier Catering Guidance for Different Types of Businesses (PHE, 2019) |
| Place pizzas directly onto the tray without greasing, use a little flour instead | site visit / questionnaire | Y/N? | Fats | Takeaway / Restaurant / Café | Y | Healthier Catering Guidance for Different Types of Businesses (PHE, 2019) |
| Use skimmed, 1% fat or semi-skimmed milk as default | site visit / questionnaire | Y/N? | Dairy/alternatives | Takeaway / Restaurant / Café | Y | Healthier Catering Guidance for Different Types of Businesses (PHE, 2019) |
| Always include at least one portion (80g) of vegetables as part of a meal | Menu analysis | Y/N? | Vegetables | Takeaway / Restaurant / Café | Y | Healthier Catering Guidance for Different Types of Businesses (PHE, 2019) |
| Meal deals or special offers should include a carbohydrate (with no added fat), vegetables and salad, and one portion of fruit | Menu analysis | Y/N? | Special offers | Takeaway / Restaurant / Café | Y | Healthier Catering Guidance for Different Types of Businesses (PHE, 2019) |
| Vegetables and starchy foods cooked without salt | site visit / questionnaire | Y/N? | Salt | Takeaway / Restaurant / Café | Y | Government Buying Standard for food and catering services (DEFRA, 2021) |
| Salt not available on tables | site visit / questionnaire | Y/N? | Salt | Takeaway / Restaurant / Café | Y | Government Buying Standard for food and catering services (DEFRA, 2021) |
| At least 75% of meat products, breads, soups, cooking sauces, ready meals, breakfast cereals, pre-packaged sandwiches should meet current core salt targets | site visit / questionnaire | Y/N? | Salt | Takeaway / Restaurant / Café | Y | Government Buying Standard for food and catering services (DEFRA, 2021) |
| A portion of fruit should be cheaper than a portion of hot or cold dessert | Menu analysis / site visit / questionnaire | Y/N? | Fruit | Takeaway / Restaurant / Café | Y | Government Buying Standard for food and catering services (DEFRA, 2021) |
| Half of desserts available should be at least 50% of the weight as fruit | Menu analysis / site visit / questionnaire | Y/N? | Fruit / Desserts | Takeaway / Restaurant / Café | Y | Government Buying Standard for food and catering services (DEFRA, 2021) |
| Main meals within a meal deal should include a starchy carbohydrate (not prepared with fats or oils), at least 1 portion of vegetables and 1 portion of fruit | Menu analysis | Y/N? | Special offers | Takeaway / Restaurant / Café | Y | Government Buying Standard for food and catering services (DEFRA, 2021) |
| Any foods and drinks within a meal deal must meet relevant GBSF standards for healthier options | Menu analysis / site visit / questionnaire | Y/N? | Special offers | Takeaway / Restaurant / Café | Y | Government Buying Standard for food and catering services (DEFRA, 2021) |
| At least 50% of hard yellow cheese procured by volume shall have a maximum total fat content of 25g/100g | site visit / questionnaire | Y/N? | Fats | Takeaway / Restaurant / Café | Y | Government Buying Standard for food and catering services (DEFRA, 2021) |
| At least 75% of ready meals procured by volume shall contain less than 6g saturated fat per portion | site visit / questionnaire | Y/N? | Fats | Takeaway / Restaurant / Café | Y | Government Buying Standard for food and catering services (DEFRA, 2021) |
| At least 75% of milk procured by volume is lower fat (semi-skimmed, 1% or skimmed milk) | site visit / questionnaire | Y/N? | Dairy/alternatives | Takeaway / Restaurant / Café | Y | Government Buying Standard for food and catering services (DEFRA, 2021) |
| At least 75% of oils and 75% of spreads procured by volume are based on unsaturated fats | site visit / questionnaire | Y/N? | Fats | Takeaway / Restaurant / Café | Y | Government Buying Standard for food and catering services (DEFRA, 2021) |
| At least 50% of pre-packed sandwiches and other savoury pre-packed meals (wraps, salads, pasta salads) provided contain 400kcal (1680 kJ) or less per serving and do not exceed 5.0g saturated fat per 100g | site visit / questionnaire | Y/N? | Healthy / healthier options | Takeaway / Restaurant / Café | Y | Government Buying Standard for food and catering services (DEFRA, 2021) |
| At least 50% of bread provided contains at least 3g fibre per 100g excluding pre-packed sandwiches | site visit / questionnaire | Y/N? | Grains / fibre | Takeaway / Restaurant / Café | Y | Government Buying Standard for food and catering services (DEFRA, 2021) |
| At least 75% of pre-packed sandwiches provided contains bread with at least 3g fibre per 100g | site visit / questionnaire | Y/N? | Grains / fibre | Takeaway / Restaurant / Café | Y | Government Buying Standard for food and catering services (DEFRA, 2021) |
| At least 75% of products provided that are included in the following categories covered by the sugar reduction programme to not exceed the following: biscuits - 100kcal, cakes - 220 kcal, morning goods - 220 kcal, puddings - 220 kcal, ice cream - 220 kcal, yogurts - 120 kcal | site visit / questionnaire | Y/N? | Sugar | Takeaway / Restaurant / Café | Y | Government Buying Standard for food and catering services (DEFRA, 2021) |
| At least 50% of breakfast cereals provided are higher in fibre (that is, at least 6g/100g) and shall not exceed 12.3g/100g total sugars (10g additional allowance for dried fruit in cereal) | site visit / questionnaire | Y/N? | Healthy / healthier options | Takeaway / Restaurant / Café | Y | Government Buying Standard for food and catering services (DEFRA, 2021) |
| If caterers serve lunch and an evening meal, fish is provided twice per week (2X 140g portion), one of which is oily. If caterers only serve lunch or an evening meal, oily fish (140g portion) is available at least once every three weeks | Menu analysis / site visit / questionnaire | Y/N? | Meat/alternatives | Takeaway / Restaurant / Café | Y | Government Buying Standard for food and catering services (DEFRA, 2021) |
| Savoury snacks are only provided in packet sizes of 35g or less | Menu analysis / site visit / questionnaire | Y/N? | Smaller portions | Takeaway / Restaurant / Café | Y | Government Buying Standard for food and catering services (DEFRA, 2021) |
| At least 75% of confectionery and packet sweet snacks provided are in the smallest standard single serve portion size available within the market and do not exceed 200 kcal (maximum) for chocolate and 125 kcal (maximum) for sugar confectionery | Menu analysis / site visit / questionnaire | Y/N? | Smaller portions | Takeaway / Restaurant / Café | Y | Government Buying Standard for food and catering services (DEFRA, 2021) |
| No more than 10% beverages provided can be sugar sweetened beverages (SSB) | Menu analysis / site visit / questionnaire | Y/N? | Sugar-sweetened beverages | Takeaway / Restaurant / Café | Y | Government Buying Standard for food and catering services (DEFRA, 2021) |
| At least 90% of beverages provided must be low calorie/no added sugar beverages | Menu analysis / site visit / questionnaire | Y/N? | Diet drinks | Takeaway / Restaurant / Café | Y | Government Buying Standard for food and catering services (DEFRA, 2021) |
| All SSB to be no more than 330ml pack size. Any SSB that are hot or cold milk-based drinks including milk substitute drinks such as soya, almond, hemp, oat, hazelnut or rice need to meet 300kcal cap | Menu analysis / site visit / questionnaire | Y/N? | Sugar-sweetened beverages | Takeaway / Restaurant / Café | Y | Government Buying Standard for food and catering services (DEFRA, 2021) |
| Any meal deals should not include any SSB | Menu analysis / site visit / questionnaire | Y/N? | Special offers | Takeaway / Restaurant / Café | Y | Government Buying Standard for food and catering services (DEFRA, 2021) |
| At least 75% fruit juice, vegetable juice and smoothies to be provided in single serve packs | Menu analysis / site visit / questionnaire | Y/N? | Smaller portions | Takeaway / Restaurant / Café | Y | Government Buying Standard for food and catering services (DEFRA, 2021) |
| Maximum calories per main meal 1345kcal | lab analysis / source meal components in nutrition database | Count number | Smaller portions | Takeaway / Restaurant / Café | Y | Calorie reduction. Technical report: guidelines for industry, 2017 baseline calorie levels and the next steps (PHE, 2020) |
| Maximum calories per starter 600kcal | lab analysis / source meal components in nutrition database | Count number | Smaller portions | Takeaway / Restaurant / Café | Y | Calorie reduction. Technical report: guidelines for industry, 2017 baseline calorie levels and the next steps (PHE, 2020) |
| Maximum calories per children's meal bundle 825kcal | lab analysis / source meal components in nutrition database | Count number | Smaller portions | Takeaway / Restaurant / Café | Y | Calorie reduction. Technical report: guidelines for industry, 2017 baseline calorie levels and the next steps (PHE, 2020) |
| Maximum calories per pizza 1230kcal | lab analysis / source meal components in nutrition database | Count number | Smaller portions | Takeaway / Restaurant / Café | Y | Calorie reduction. Technical report: guidelines for industry, 2017 baseline calorie levels and the next steps (PHE, 2020) |
| Maximum calories per pastry product 670kcal | lab analysis / source meal components in nutrition database | Count number | Smaller portions | Takeaway / Restaurant / Café | Y | Calorie reduction. Technical report: guidelines for industry, 2017 baseline calorie levels and the next steps (PHE, 2020) |
| Maximum calories per sandwich 580kcal | lab analysis / source meal components in nutrition database | Count number | Smaller portions | Takeaway / Restaurant / Café | Y | Calorie reduction. Technical report: guidelines for industry, 2017 baseline calorie levels and the next steps (PHE, 2020) |
| No price promotions of HFSS food and drink | Menu analysis | Y/N? | Special offers | Takeaway / Restaurant / Café | Y | Recipe for health: a plan to fix our broken food system. (House of Lords. Food, Diet and Obesity Committee, 24/10/2024) |
| No location promotions of HFSS food and drink | site visit / questionnaire | Y/N? | Positional promotions | Takeaway / Restaurant / Café | Y | Recipe for health: a plan to fix our broken food system. (House of Lords. Food, Diet and |
|  |  |  |  |  |  | Obesity Committee,<br>24/10/2024) |
| What healthier options are available? These options must be lower in fat, salt, and/or sugar than standard menu options. | Menu analysis |  | Healthy / healthier options | Takeaway / Restaurant / Café | Y | Recipe 4 Health<br>(Lancashire County Council) |
| Where are healthier options shown in the menu and how are they priced in comparison to standard options? Please provide details AND a copy of your menu. | Menu analysis |  | Healthy / healthier options | Takeaway / Restaurant / Café | Y | Recipe 4 Health<br>(Lancashire County Council) |
| Do you use trimmed meat and poultry where possible? | site visit / questionnaire |  | Healthy / healthier options | Takeaway / Restaurant / Café | Y | Recipe 4 Health<br>(Lancashire County Council) |
| Do you use reduced fat alternatives of cream and milk in cooking? Are these available as options for serving? **Please provide product details** | site visit / questionnaire |  | fats and frying | Takeaway / Restaurant / Café | Y | Recipe 4 Health<br>(Lancashire County Council) |
| Do you grill or steam any menu items in preference to frying? **Please give details** | site visit / questionnaire |  | fats and frying | Takeaway / Restaurant / Café | Y | Recipe 4 Health<br>(Lancashire County Council) |
| Do you shallow fry menu items instead of deep frying? **Please give details** | site visit / questionnaire |  | fats and frying | Takeaway / Restaurant / Café | Y | Recipe 4 Health<br>(Lancashire County Council) |
| If processed products such as soups, sauces and pies are bought in, what lower salt and/or sugar options do you provide? **Please give examples.** | site visit / questionnaire |  | Healthy / healthier options | Takeaway / Restaurant / Café | Y | Recipe 4 Health<br>(Lancashire County Council) |
| Do you offer tap water and free water refills? | site visit / questionnaire |  | Water | Takeaway / Restaurant / Café | Y | Recipe 4 Health<br>(Lancashire County Council) |
| Are vegetables or salad included as an accompaniment as standard and are they incorporated into dishes wherever possible? **Please give examples** | Menu analysis |  | vegetables / salad | Takeaway / Restaurant / Café | Y | Recipe 4 Health<br>(Lancashire County Council) |
| Are staff knowledgeable on how to choose a healthier diet and are they able to advise customers on healthier options? **Please give details** | site visit / questionnaire |  | staff | Takeaway / Restaurant / Café | Y | Recipe 4 Health<br>(Lancashire County Council) |
| Is there an oily fish option available e.g. tuna, salmon, mackerel, pilchards, and sardines. **Please give details** NB tuna is NOT classed as an oily fish when canned but the others are | Menu analysis |  | Healthy / healthier options | Takeaway / Restaurant / Café | Y | Recipe 4 Health<br>(Lancashire County Council) |
| Are sandwiches served without spread as standard unless the customer specifically requests it? | site visit / questionnaire |  | fats | Takeaway / Restaurant / Café | Y | Recipe 4 Health<br>(Lancashire County Council) |
| Are fruit and fruit-based desserts available? e.g. whole fresh fruit, fruit salads, malt loaf, fruit cake, fruit crumbles | Menu analysis / site visit / questionnaire |  | fruit | Takeaway / Restaurant / Café | Y | Recipe 4 Health<br>(Lancashire County Council) |
| Do you provide low fat sauces, or do you prepare your own sauces with low fat ingredients? **Please give details** | site visit / questionnaire |  | fats | Takeaway / Restaurant / Café | Y | Recipe 4 Health (Lancashire County Council) |
| What type of oils do you use? Unsaturated oils and fats are better e.g. olive oil, rapeseed, sunflower oil, non-blended vegetable oils, groundnut oil | site visit / questionnaire |  | fats | Takeaway / Restaurant / Café | Y | Recipe 4 Health (Lancashire County Council) |
| If you offer fried fish, is this lightly battered? | site visit / questionnaire |  | fats and frying | Takeaway / Restaurant / Café | Y | Recipe 4 Health (Lancashire County Council) |
| Do you offer baked potatoes with low fat fillings? (e.g., cottage cheese, tuna with low fat mayo, beans) **Please give examples** | Menu analysis |  | Healthy / healthier options | Takeaway / Restaurant / Café | Y | Recipe 4 Health (Lancashire County Council) |
| Do you provide thick cut chips or potato wedges as an alternative to French fries/ thin chips? | Menu analysis / site visit / questionnaire |  | chips | Takeaway / Restaurant / Café | Y | Recipe 4 Health (Lancashire County Council) |
| What bread options are available? e.g. 50/50, wholemeal, granary | Menu analysis |  | Grains / fibre | Takeaway / Restaurant / Café | Y | Recipe 4 Health (Lancashire County Council) |
| Do you provide low fat salad dressings? **Please give details** | Menu analysis / site visit / questionnaire |  | fats | Takeaway / Restaurant / Café | Y | Recipe 4 Health (Lancashire County Council) |
| What healthier drinks are available? e.g. water, pure fruit juice, semi and/or skimmed milk drinks | Menu analysis |  | Drinks | Takeaway / Restaurant / Café | Y | Recipe 4 Health (Lancashire County Council) |
| If making homemade chips, do you keep the skin on or try to source chips with the skin on? | Menu analysis / site visit / questionnaire |  | chips | Takeaway / Restaurant / Café | Y | Recipe 4 Health (Lancashire County Council) |
| What fruits and vegetables do you have available as snacks? (If appropriate for your cuisine) | Menu analysis / site visit / questionnaire |  | Fruit and vegetables | Takeaway / Restaurant / Café | Y | Recipe 4 Health (Lancashire County Council) |
| Do you serve one portion of vegetables in each meal? (Around 80g of vegetables) | Menu analysis / site visit / questionnaire |  | Vegetables | Takeaway / Restaurant / Café | Y | Recipe 4 Health (Lancashire County Council) |
| Do you use reduced hole saltshakers? i.e. with 3 holes or less, rather than 5 or more | site visit / questionnaire |  | salt | Takeaway / Restaurant / Café | Y | Recipe 4 Health (Lancashire County Council) |
| How is salt minimised in recipes? Please provide details | site visit / questionnaire |  | salt | Takeaway / Restaurant / Café | Y | Recipe 4 Health (Lancashire County Council) |
| Do you source low-salt stock cubes, gravy granules and sauces (where applicable) **Please provide product details** | site visit / questionnaire |  | salt | Takeaway / Restaurant / Café | Y | Recipe 4 Health (Lancashire County Council) |
| Do you reduce or refrain from using MSG (Monosodium Glutamate) AND salt together in recipes? | site visit / questionnaire |  | salt | Takeaway / Restaurant / Café | Y | Recipe 4 Health (Lancashire County Council) |
| Do you avoid adding salt when boiling potatoes, rice, or pasta? | site visit / questionnaire |  | salt | Takeaway / Restaurant / Café | Y | Recipe 4 Health (Lancashire County Council) |
| Do you refrain from serving unlimited refills of sugar sweetened drinks? | site visit / questionnaire |  | Sugar-sweetened beverages | Takeaway / Restaurant / Café | Y | Recipe 4 Health (Lancashire County Council) |
| Where hot drinks are served, including self-service, do you offer sweeteners as an alternative to sugar? | site visit / questionnaire |  | drinks | Takeaway / Restaurant / Café | Y | Recipe 4 Health (Lancashire County Council) |
| Are all fruit juices pure and unsweetened and available in 150ml servings? Please provide product details | Menu analysis / site visit / questionnaire |  | fruit juice | Takeaway / Restaurant / Café | Y | Recipe 4 Health (Lancashire County Council) |
| Do you ensure that products do not contain hydrogenated fats? **Please explain how do you do this?** | site visit / questionnaire |  | fats | Takeaway / Restaurant / Café | Y | Recipe 4 Health (Lancashire County Council) |
| Do you consider where and how Trans Fatty Acids can be reduced or eliminated from the menu? e.g. only purchasing products that are free from Trans Fatty Acids | site visit / questionnaire |  | fats | Takeaway / Restaurant / Café | Y | Recipe 4 Health (Lancashire County Council) |
| Which types of milk do you use or serve as standard for hot drinks? i.e. not using whole milk as standard (exceptions for early years settings) or verbally offering semi or skimmed milk | site visit / questionnaire |  | Dairy/alternatives | Takeaway / Restaurant / Café | Y | Recipe 4 Health (Lancashire County Council) |
| Do you operate the 'bang, shake and drain' method for frying to help reduce the amount of oil absorbed? | site visit / questionnaire |  | fats and frying | Takeaway / Restaurant / Café | Y | Recipe 4 Health (Lancashire County Council) |
| Do you look at oil maintenance and how this could be improved (e.g. oil in deep fryers is regularly strained/debris skimmed after each batch/throughout service, fully filtered at least weekly, oil is topped up after every session and oil is changed before it froths, foams or smokes) Please state what you do | site visit / questionnaire |  | fats and frying | Takeaway / Restaurant / Café | Y | Recipe 4 Health (Lancashire County Council) |
| Please detail any healthier products you promote or market. | Menu analysis / site visit / questionnaire |  | Healthy / healthier options | Takeaway / Restaurant / Café | Y | Recipe 4 Health (Lancashire County Council) |
| How do you promote healthier options? **Please explain** | Menu analysis / site visit / questionnaire |  | Healthy promotions | Takeaway / Restaurant / Café | Y | Recipe 4 Health (Lancashire County Council) |
| Do you promote healthy eating guidelines on your premises? e.g. 5-a-day, the Eatwell plate, reduced salt intake. **Please give details** | site visit / questionnaire |  | Healthy promotions | Takeaway / Restaurant / Café | Y | Recipe 4 Health (Lancashire County Council) |
| Do you keep food or drinks high in fat, sugar, or salt away from the point of purchase? i.e. at the serving counter / till point | site visit / questionnaire |  | Positional promotions | Takeaway / Restaurant / Café | Y | Recipe 4 Health (Lancashire County Council) |
| Do you avoid prompting customers to take larger servings? e.g. 'extra toppings' or 'go large' | site visit / questionnaire |  | Overconsumption | Takeaway / Restaurant / Café | Y | Recipe 4 Health (Lancashire County Council) |

**Supplementary Material 2.**
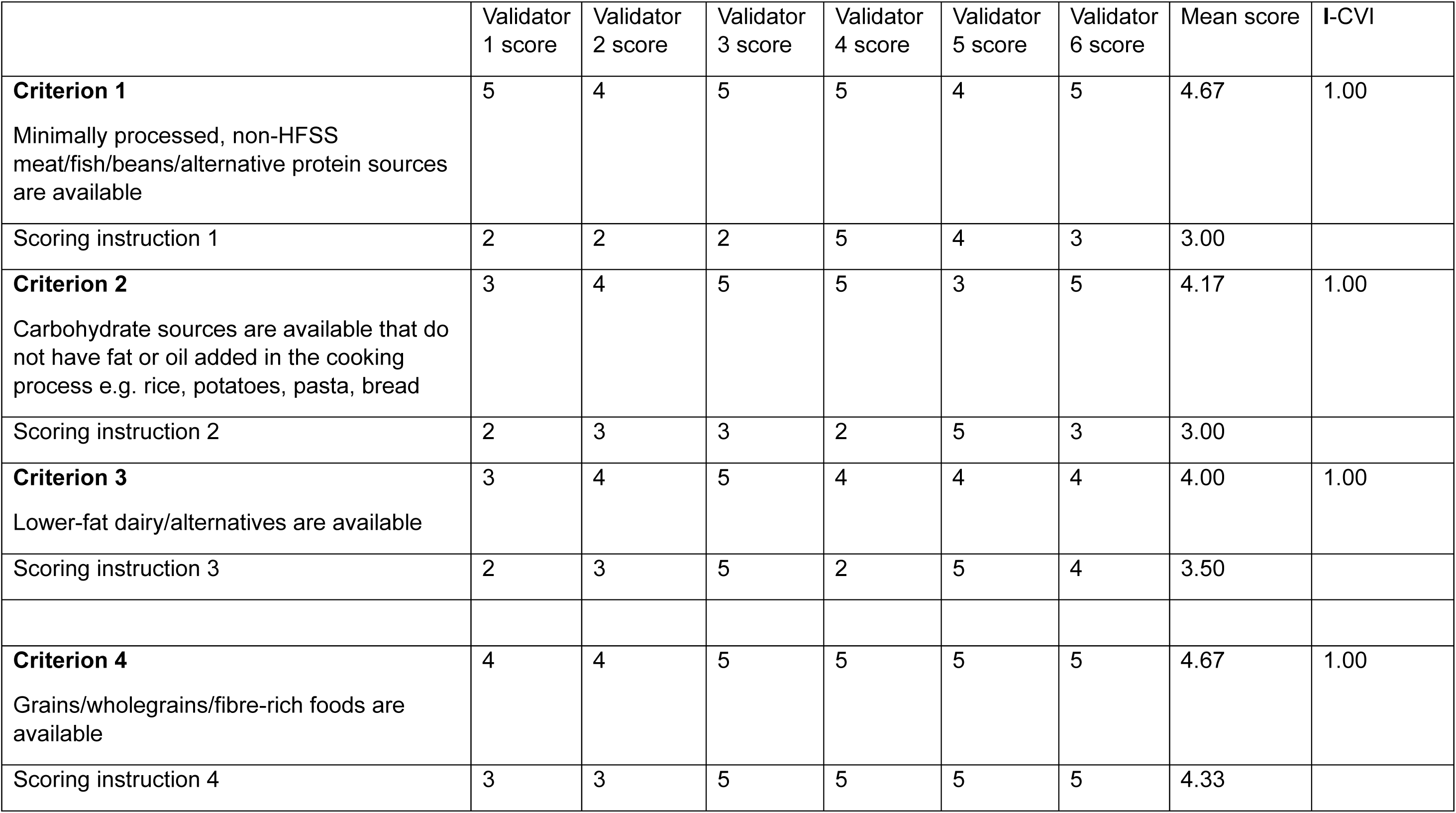
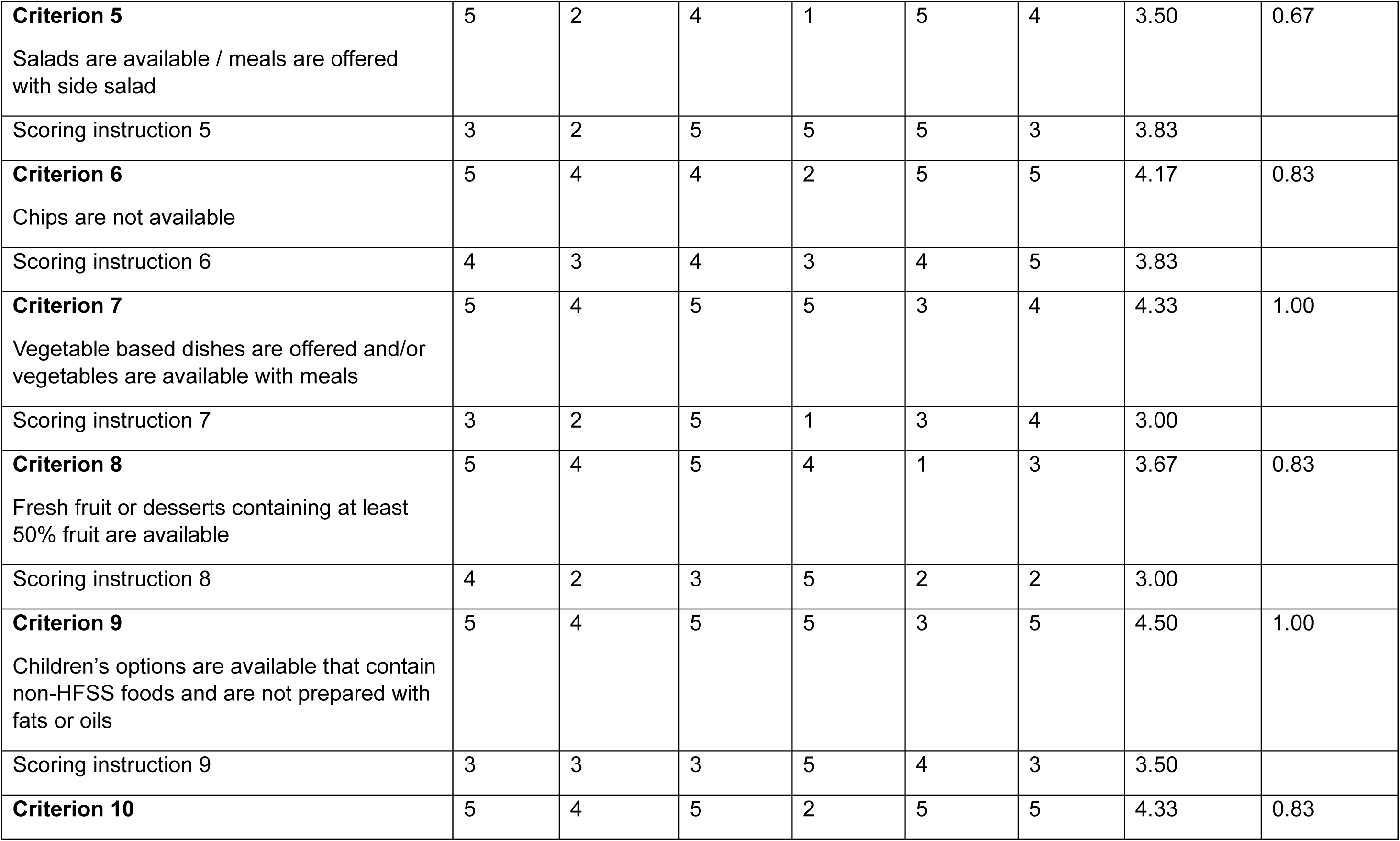
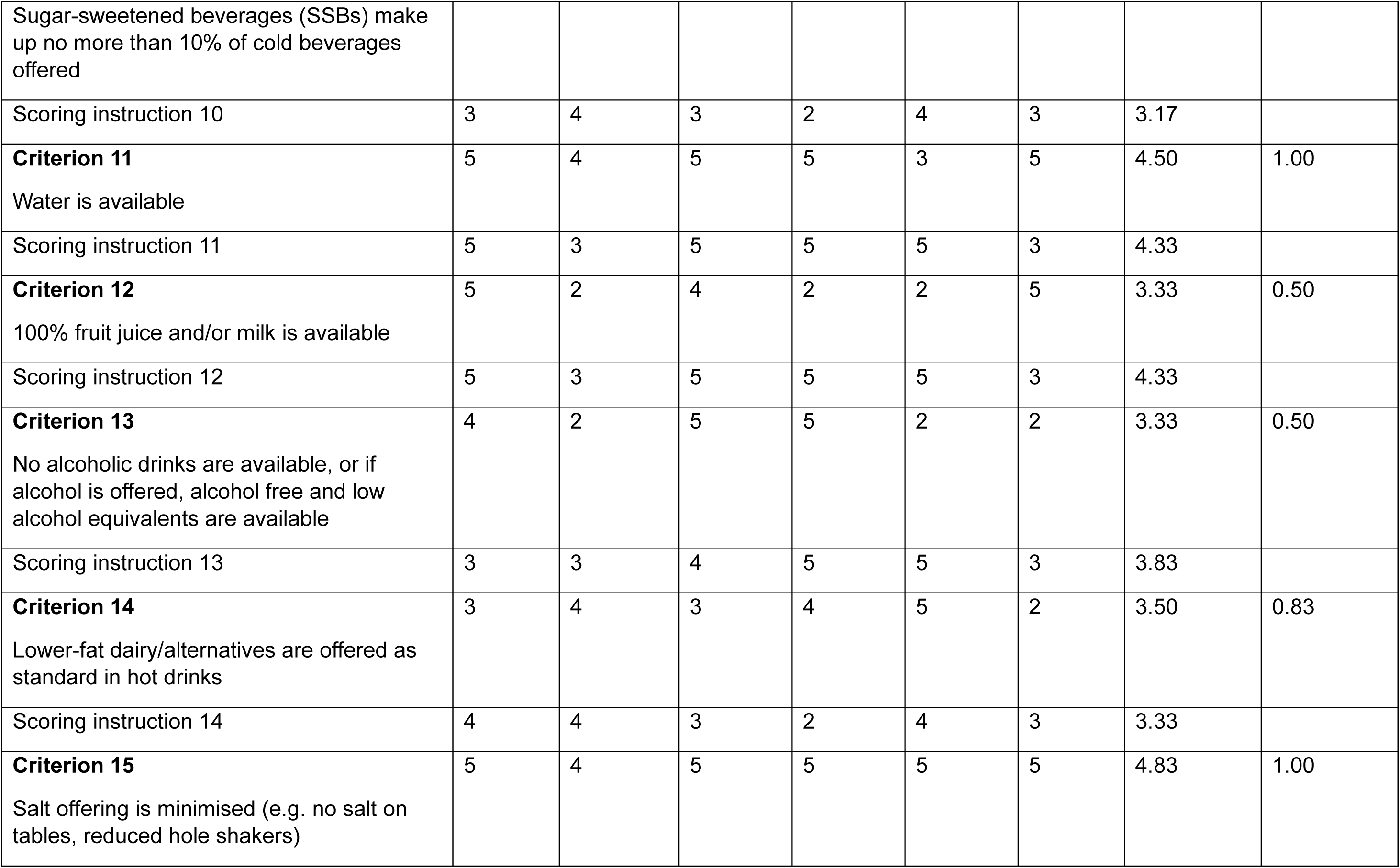
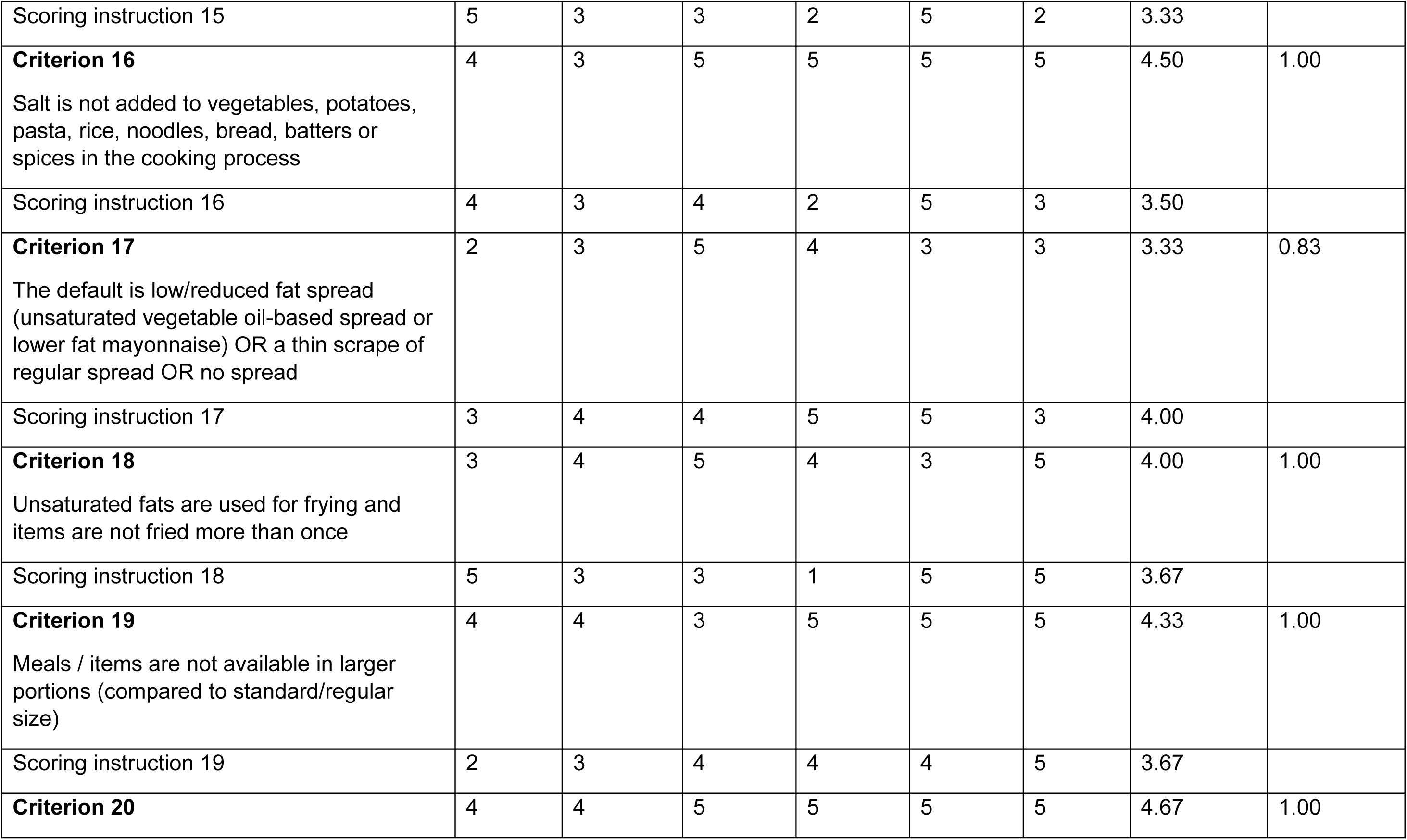
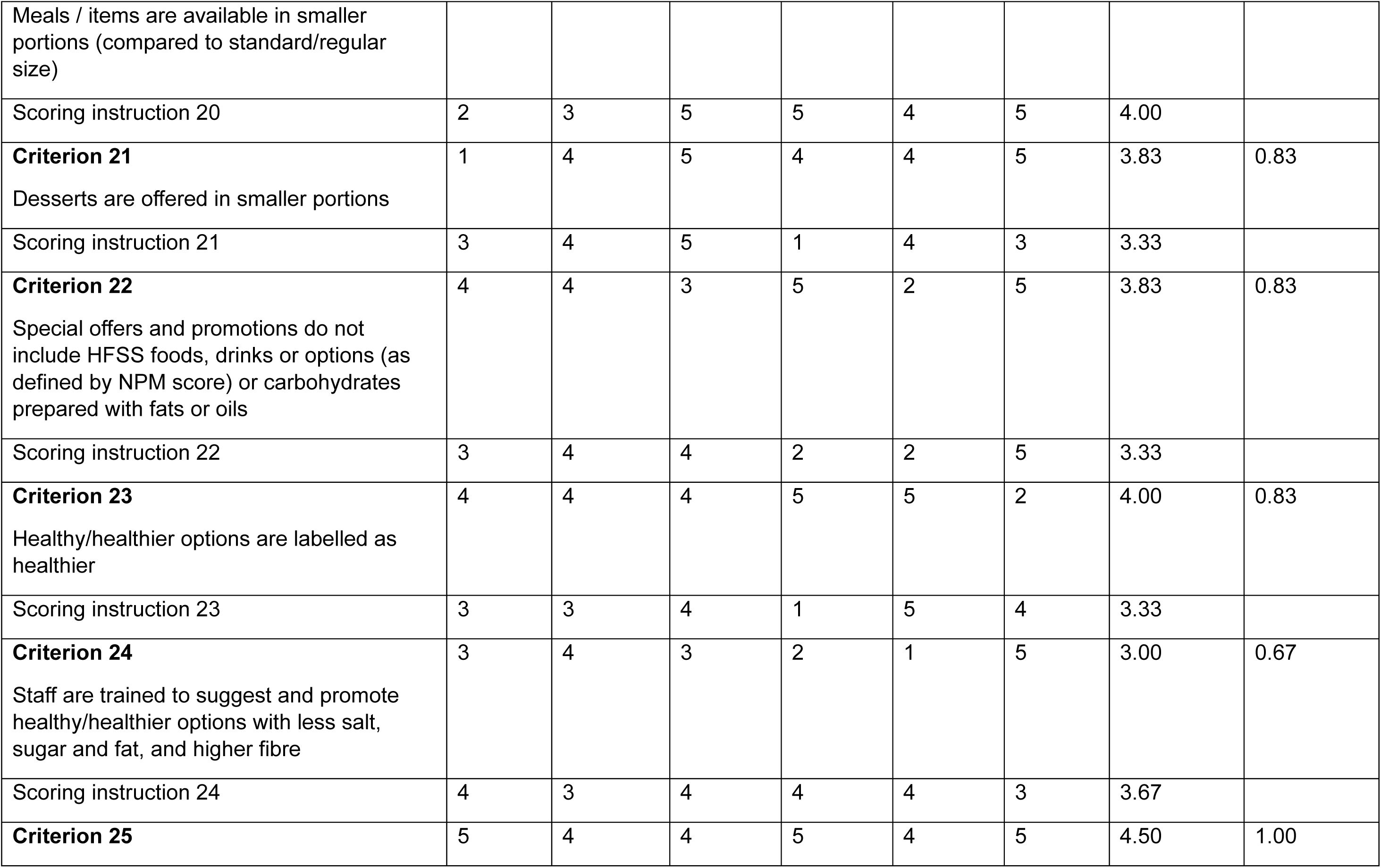
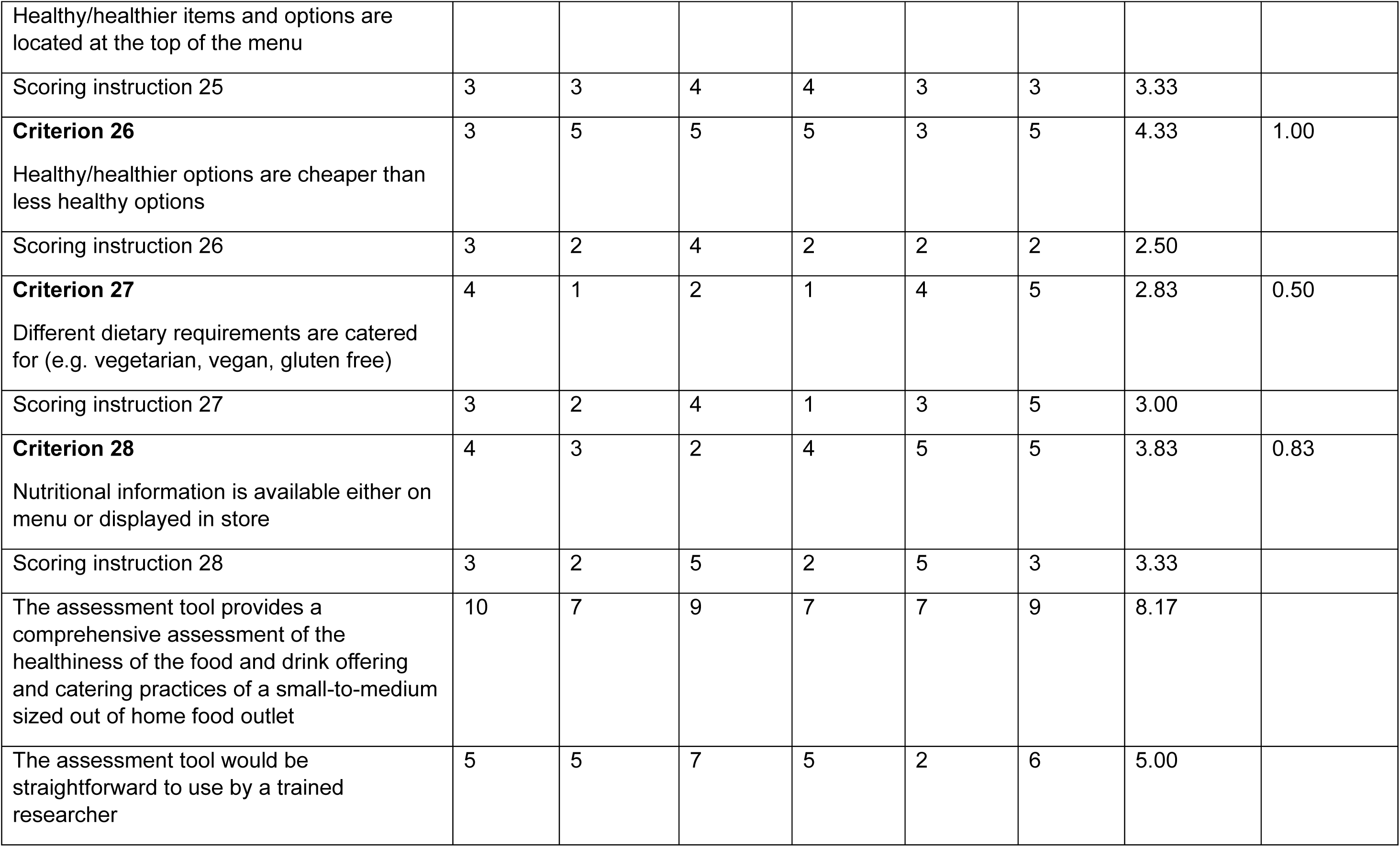
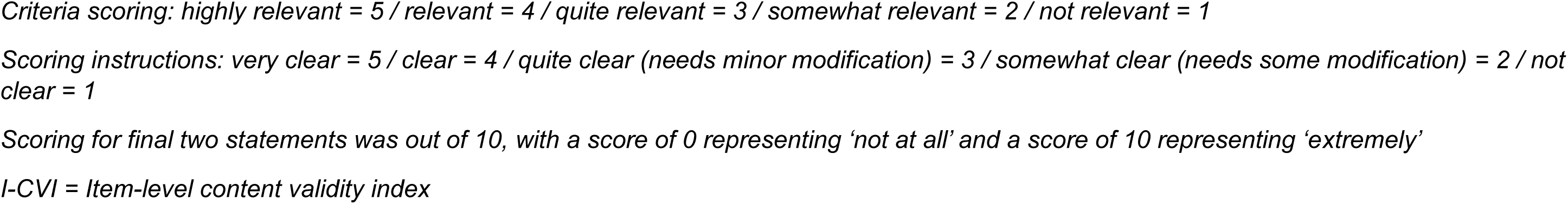
Validator scores from content validity assessment.

**Supplementary Material 3.** OHFS menu healthiness assessment tool (Menu-Health)

| Criterion number | Category | Menu (A) or contact needed (B) | Criterion | Scoring instructions | Further instructions / examples / resources | enter business name here | enter business name here |
| --- | --- | --- | --- | --- | --- | --- | --- |
| 1 | Healthier meal components | A | Non HFSS protein sources (meat/fish/beans/alternative proteins) are available that are not deep fried and are prepared without with large amounts of fat or oil | Non HFSS protein sources (meat/fish/beans/alternative proteins) are available that are not deep fried and not prepared with large amounts of fat or oil | For 'dessert only' businesses select N/A. https://www.gov.uk/government/publications/the-eatwell-guide. Examples of acceptable protein sources include: grilled, baked, boiled or shallow fried chicken, fish, shellfish, beef, pork, lamb, mince, tofu, tempeh, beans, chickpeas, lentils, soy protein, soya beans. Composite dishes including a non HFSS protein source also count, e.g. spaghetti bolognese, fish pie, chickpea curry, griddled chicken kebab. Exclusions: deep fried meat and fish, donner kebabs, burgers, processed meats (e.g. sausages, salami, ham, bacon) | 0 | 0 |
| 2 | Healthier meal components | A | Carbohydrate sources are available that are not deep fried and are prepared without large amounts of fat or oil | Carbohydrate sources are available that are not deep fried or do not have large amounts of fat or oil added in the cooking process | For 'dessert only' businesses select N/A. Eligible items can include a drizzle of oil or small amount of butter/fat e.g. naan bread, garlic naan, fried rice. Exclusions: deep fried chips, deep fried potatoes, sweet potatoes fries, roast potatoes, fried potatoes, fried bread and croutons, deep fried noodles, garlic bread, pizza, | 0 | 0 |
| 3 | Healthier meal components | A | Lower-fat dairy/alternatives are available | Score of 1 allocated if any of the dairy products / dairy alternatives offered or used in cooking are lower-fat options. Score of zero allocated if no lower-fat dairy products or alternatives are offered / used. | Examples of lower-fat dairy products: Cottage cheese, quark, reduced / lower fat hard cheese, semi-skimmed and skimmed milk, lower/reduced fat yoghurt. If there is no information about the dairy used, select 'no data on menu'. If no dairy products are used or offered select N/A. | 0 | 0 |
| 4 | Healthier meal components | A | Grains / wholegrains / fibre rich carbohydrates are available | Score of 1 allocated if any of the carbohydrate sources offered are considered a source of fibre (3g fibre/100g food) or high fibre (6g fibre/100g food). Score of zero allocated if no carbohydrates considered a source of fibre / high fibre carbohydrates are offered. | Examples of higher fibre carbohydrates: wholemeal bread, granary bread, brown bread, seeded bread, wholewheat couscous, quinoa, wholewheat pasta, brown rice, roti/chapati, oats. If there is no information about the carbohydrate sources used, select 'no data on menu'. | 0 | 0 |
| 5 | Healthier meal components | A | Vegetable predominant salads are available / meals are offered with side salad | Score of 1 allocated if any salads comprised predominantly of vegetables are listed on the menu or if any items come with a side salad (not including small salad garnish). Score of zero allocated if no salads / side salads available. | Salad would be based around cold items e.g. lettuce/leaves, tomato, cucumber, peppers, onion, carrot, coleslaw. Excluded salads would be those that contain large amounts of HFSS items (by NPM score), and large amounts of higher-fat salad dressing. For cuisines that do not sell salad (e.g. Chinese) and 'dessert only' businesses select N/A. | 0 | 0 |
| 6 | Healthier meal components | A | Deep-fried chips are not available | Score of 1 allocated if deep-fried chips or deep-fried sweet potato fries are not available (can offer baked / air fried / wedges as long as not deep fried). Score of zero allocated if chips / fries / sweet potato fries are available. | For 'dessert only' businesses select N/A. | 0 | 0 |
| 7 | Healthier meal components | A | Vegetable based dishes are offered and/or vegetables are available with meals | Score of 1 allocated if any vegetable based dishes (containing at least one portion of vegetables) are listed on the menu or if any items come with a portion of vegetables as standard. Score of zero allocated if no vegetables / vegetable based dishes available. | Vegetable accompaniments would typically be served hot (compared to side salad which would be cold). Examples include: beans, legumes and pulses (e.g. baked beans, soybeans, cannellini beans, chickpeas etc) and vegetable based soups but not potatoes. For 'dessert only' businesses select N/A. Examples of vegetable based dishes: bean chilli, vegetable stir fry, lentil curry, cauliflower steak | 0 | 0 |
| 8 | Healthier meal components | A | Fresh fruit is available | Score of 1 allocated if fresh fruit / fruit salad is available. | Can include fruit offered for children and fruit salad. If no desserts (and therefore no fruit) listed on menu, select N/A. | 0 | 0 |
| 9 | Healthier meal components | A | Children's options are available that contain non-HFSS foods and are prepared without large amounts of fat or oil | Score of 1 allocated if there is at least one children's meal option that does not contain HFSS items (as per NPM score) or items prepared with large amounts of fats or oils. Score of zero allocated if all children's meal options contain HFSS foods / are prepared with large amounts of fat or oil | If unsure use NPM calculator to assess if item would be categorised as HFSS. https://npmcalculator.cdrc.ac.uk/. If nutritional information is unavailable use McCance and Widdowson nutritional database to find product most similar to that offered https://www.gov.uk/government/publications/composition-of-foods-integrated-dataset-cofid. N.B. breaded fishfingers would pass NPM scoring system (unless deep fried). Examples of acceptable meals: pasta with tomato based sauce, spaghetti bolognese, jacket potato with beans / tuna, wraps or sandwiches with non-HFSS fillings and with veg sticks, fruit, yoghurts. If no children's meals on menu select 'N/A'. | 0 | 0 |
| 10 | Healthier meal components | A | Desserts are not sold, or if desserts are sold, they are available in smaller sized portions alongside standard sized portions | Score of 1 allocated if no desserts are available. Score of 1 also allocated if desserts are sold they are also available in smaller portions alongside standard sizes. Score of zero allocated if desserts are available and not offered in smaller portions. | For 'dessert only' businesses: if smaller portions of desserts are available, score 1 point for this criterion but no additional point is allowed for criterion 21 (mark criterion 21 as N/A). | 0 | 0 |
| 11 | Healthier drinks | A | Sugar-sweetened beverages (SSBs) make up no more than 10% of cold beverages offered | Score of 1 allocated if SSBs make up no more than 10% of cold (non-alcoholic) beverages offered. Score of zero allocated if SSBs make up more than 10% of cold (non-alcoholic) beverages offered. | List all cold non-alcoholic beverages offered. Categorise as SSBs or not (categorised as SSB if drink contain at least 5g sugar per 100ml and has sugar added during production, or an ingredient that contains sugar e.g. honey (but not fruit/fruit juice or milk). Calculate 10% of SSBs of total cold beverages. Smoothies, milkshakes, frappes and iced coffees are not included in this assessment. If drinks are not listed on menu select 'no data on menu'. | 0 | 0 |
| 12 | Healthier drinks | A | Water is available | Score of 1 allocated if water is available (tap water or bottled water). Score of zero allocated if water is not available. | If drinks are not listed on menu select 'no data on menu'. Flavoured water would not be classed as water. | 0 | 0 |
| 13 | Healthier drinks | A | No alcoholic drinks are available, or if alcohol is offered, alcohol free and low alcohol equivalents are available | Score of 1 allocated if alcoholic drinks are not available. Score of 1 allocated if alcohol is offered AND alcohol free or low alcohol equivalents are also available (not including soft drinks). Score of zero allocated if alcohol free / low alcohol equivalents are not available alongside alcohol. | Examples: alcohol free beer ($\leq 0.05\%$ abv), low alcohol beer ($\leq 1.2\%$ abv), low alcohol wine ($\leq 1.2\%$ abv), dealcoholised wine ($\leq 0.5\%$ abv), alcohol free spirits. If drinks are not listed on the menu select 'no data on menu'. | 0 | 0 |
| 14 | Healthier drinks | B | Lower-fat dairy/alternatives are offered as standard in hot drinks | Score of 1 allocated if semi-skimmed or skimmed milk is used as the default option in hot drinks. Score of zero allocated if whole milk is offered as default. | If no drinks listed on menu, or if no information about milk for hot drinks on menu select 'no data on menu'. If business has cold drinks on menu but does not sell hot drinks select 'N/A'. | 0 | 0 |
| 15 | Healthier catering practices | B | Salt offering is minimised | Score of 1 allocated if any attempt is made to reduce customer salt offering. Score of zero allocated if no attempt to minimise salt availability. | Examples of attempts to reduce salt offering: no salt shakers or soy sauce on tables, customers must request salt or soy sauce, reduced hole salt shakers are used (5 holes rather than 17 in fish and chip shops and takeaways), salt is available in sachets, low-sodium salt is offered instead of salt | 0 | 0 |
| 16 | Healthier catering practices | B | Salt is not added to vegetables, pasta, rice and noodles. Minimal salt is added to potatoes, bread, and batters | Score of 1 allocated if salt is not added to vegetables, pasta, rice and noodles, and minimal salt is added to potatoes, bread or batters. Score of zero allocated if excessive / extra salt is added (e.g. large pinches added to meals when they have been cooked or plated) |  | 0 | 0 |
| 17 | Healthier catering practices | B | The default is low/reduced fat spread (unsaturated vegetable oil-based spread or lower fat mayonnaise) OR a small portion / thin scrape of butter / spread OR no spread | Score of 1 allocated if low or reduced fat spread made with unsaturated vegetable oil or a small portion / thin scrape of spread/butter or low-fat mayonnaise or no spread/butter is used as the default option on sandwiches or with jacket potatoes, bread, toast, scones. Score of zero allocated if large portion / thickly spread butter / spread / full fat mayonnaise is used as the default option. | If no information on menu select 'no data on menu'. If business does not sell sandwiches, jacket potatoes, bread (with spread/butter accompaniment), toast or scones select 'N/A'. | 0 | 0 |
| 18 | Healthier catering practices | B | Unsaturated fats are used for frying | Score of 1 allocated if unsaturated fats are always used for frying. Score of zero allocated if saturated fats are used for frying. | Examples of unsaturated fats: olive oil, rapeseed oil, sunflower oil, safflower oil, peanut oil, walnut oil, corn oil. | 0 | 0 |
| 19 | Healthier catering practices | B | Items are not fried more than once | Score of 1 allocated if all menu items are not fried more than once. Score of zero allocated if any items are fried more than once. | Examples of items fried more than once: triple cooked chips, double fried chips, some fried chicken | 0 | 0 |
| 20 | Portion size | A | Meals / items are not available in larger portions (compared to standard/regular size) | Score of 1 allocated if meals or items are not available in larger portion sizes (compared to standard / regular size). Score of zero allocated if larger meal options or larger items, 'go large' or 'add extra' are available (including crisps over 35g, SSBs over 330ml) | If unsure of which items would be classed as 'standard/regular', compare price to other similar dishes. This criterion does not apply to sharing platters / dishes intended for more than one person or hot drinks. If 'add extra/ go large' is assessed in this criterion, do not assess again as a special offer (criterion 22). | 0 | 0 |
| 21 | Portion size | A | Meals / items are available in smaller portions (compared to standard/regular size and large) | Score of 1 allocated if meals or items are available in smaller portion sizes (compared to standard / regular size, not including meals specifically marketed for children). Score of zero allocated if no smaller options are available. | If unsure of which items would be classed as 'standard/regular', compare price to other similar dishes. This criterion applies to savoury options & SSBs only and does not apply to hot drinks and desserts- assess desserts (and 'dessert only' businesses) in criterion 10 and select N/A here. | 0 | 0 |
| 22 | Special offers and promotions | A | Special offers and promotions do not include HFSS foods, drinks or options (as defined by NPM score) or items prepared with large amounts of fat or oil | Score of 1 allocated if no HFSS products or items prepared with large amounts of fat or oil are included in any promotions (e.g. special offer, meal deal, child's meal deal, price promotion, location promotion or daily special). Score of zero allocated if any item in any promotions includes any HFSS food/item prepared with large amounts of fat or oil. | Can also score 1 if there are no promotions on the menu. If nutritional information is unavailable use McCance and Widdowson nutritional database to find product most similar to that offered and NPM calculator to assess if item would be categorised as HFSS. https://npmcalculator.cdrc.ac.uk/ https://www.gov.uk/government/publications/composition-of-foods-integrated-dataset-cofid | 0 | 0 |
| 23 | Special offers and promotions | A | Healthy/healthier options are promoted on menus | Score of 1 allocated if any healthier items or meals are either labelled as healthy / healthier options on menus, in the most prominent position on the menu (e.g. at top, in a central box), or promoted by signage. Score of zero allocated if no healthy / healthier foods, items or meals are promoted. | Healthier options include those with less salt, sugar and fat, and more fibre, vegetables, salad and wholegrains- refer to Eatwell guide for guidance. | 0 | 0 |
| 24 | Special offers and promotions | A | Labelled healthy/healthier options are cheaper than less healthy options | Score of 1 allocated if labelled healthy/healthier options are cheaper than less healthy options. Score of zero allocated if healthy/healthier options are not cheaper than less healthy options. Select N/A if healthier items are not labelled. | For any labelled 'healthier' menu items, select a comparable 'less healthy' item and compare price. Healthier options include those with less salt, sugar and fat, and more fibre, vegetables, salad and wholegrains- refer to Eatwell guide / NPM calculator / McCance and Widdowson for guidance. https://npmcalculator.cdrc.ac.uk/ https://www.gov.uk/government/publications/composition-of-foods-integrated-dataset-cofid | 0 | 0 |
| 25 | Diet and nutrition information | A | Healthier options are available that cater for different dietary requirements (e.g. vegetarian or vegan or gluten free) | Score of 1 allocated if different dietary requirements are catered for (e.g. vegetarian or vegan or gluten free) AND any of these options contain non-HFSS foods (by NPM score) that are not prepared with large amounts of fat or oil | If unsure of HFSS assessment use NPM calculator / McCance and Widdowson for guidance. https://npmcalculator.cdrc.ac.uk/ https://www.gov.uk/government/publications/composition-of-foods-integrated-dataset-cofid | 0 | 0 |
| 26 | Diet and nutrition information | A | Calorie information is available on menus | Score of 1 allocated if calorie information is provided on the menu (other than the information presented on pre-packaged foods). Score of zero allocated if no calorie information is provided on the menu. |  | 0 | 0 |
|  |  |  |  |  | **Total N/A** | 0 | 0 |
|  |  |  |  |  | Total unable to assess | 0 | 0 |
|  |  |  |  |  | Number of standards assessed | 26 | 26 |
|  |  |  |  |  | Score | 0 | 0 |
|  |  |  |  |  | % healthiness | 0.00% | 0.00% |

**Supplementary Material 4.**
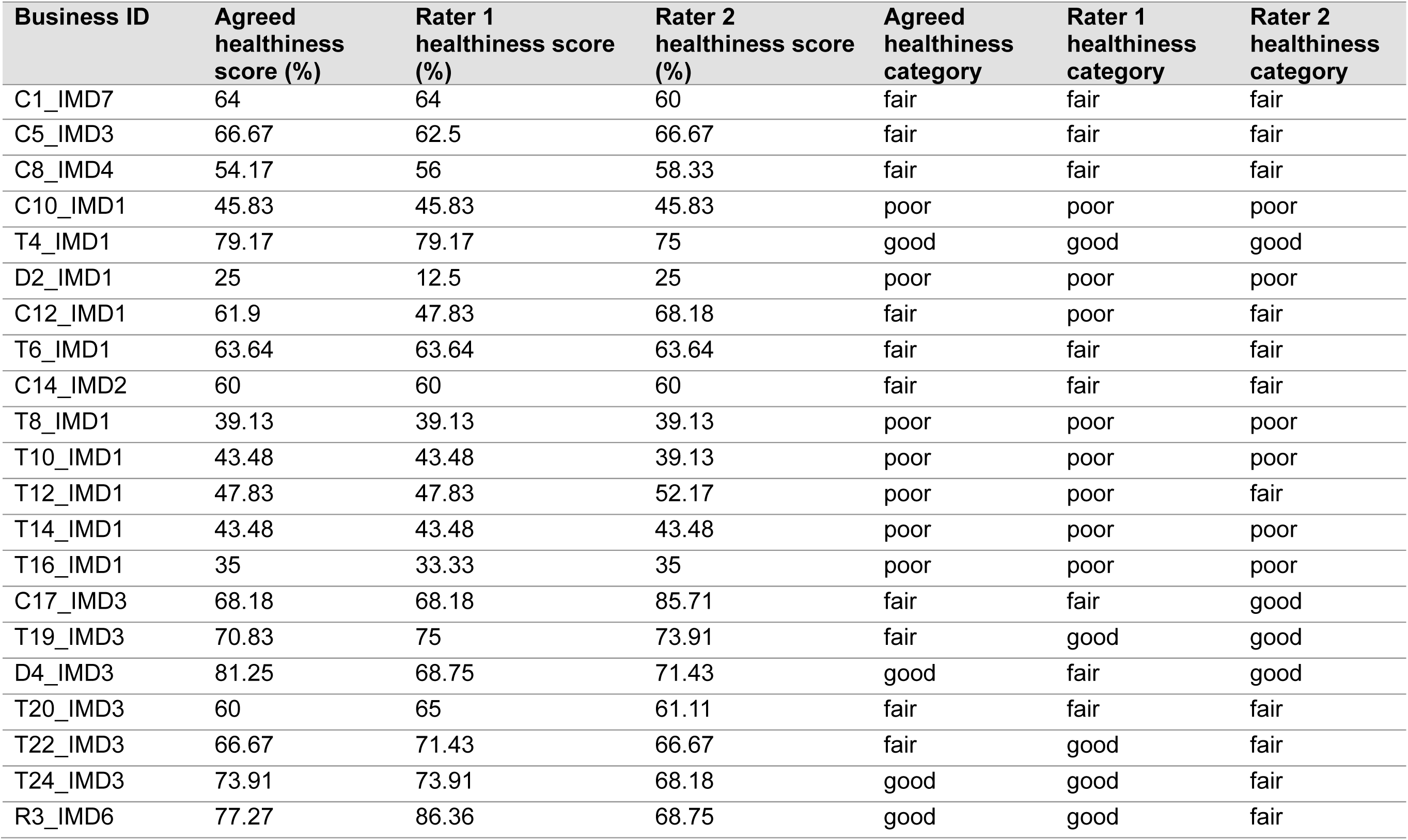
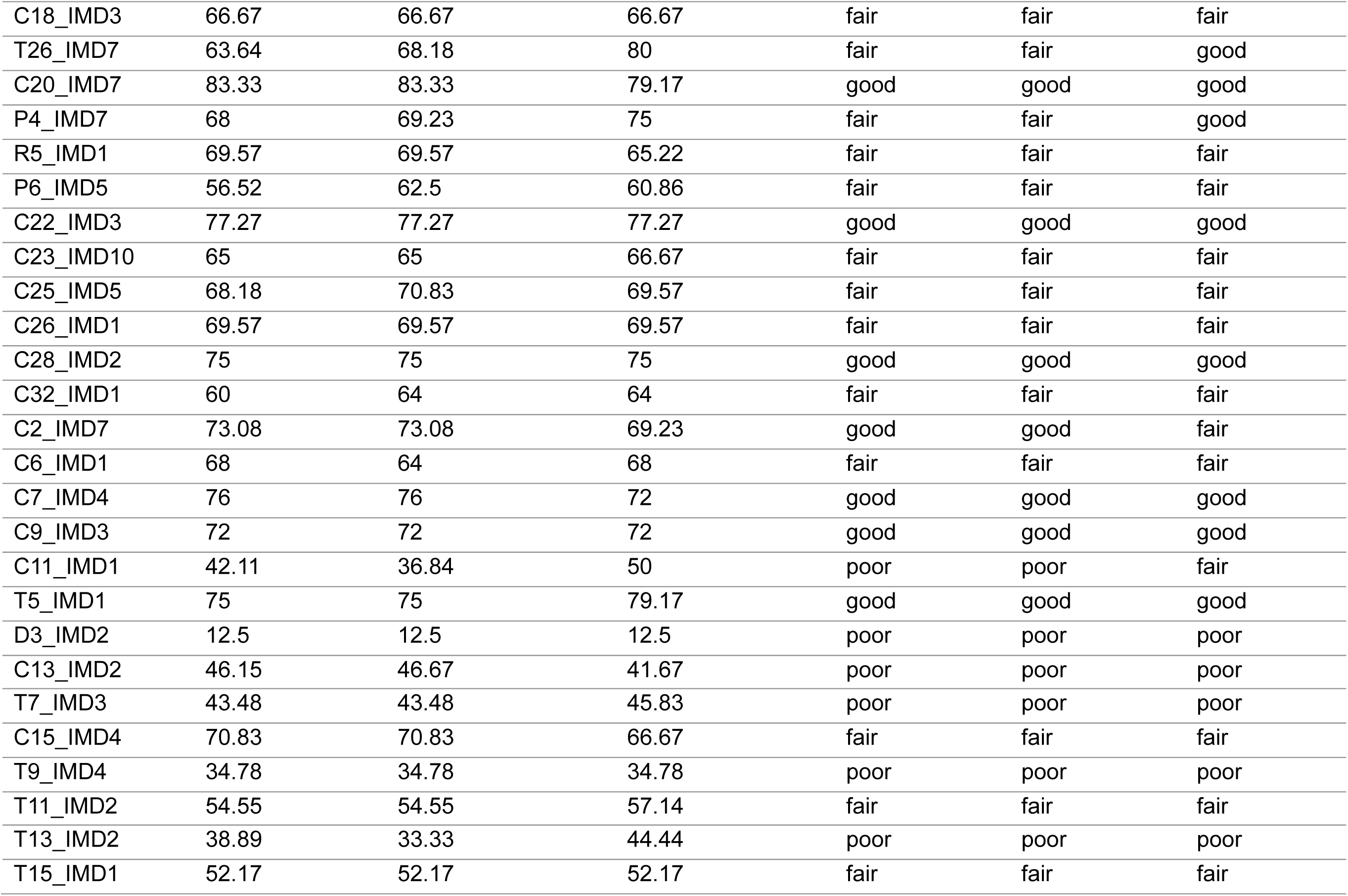
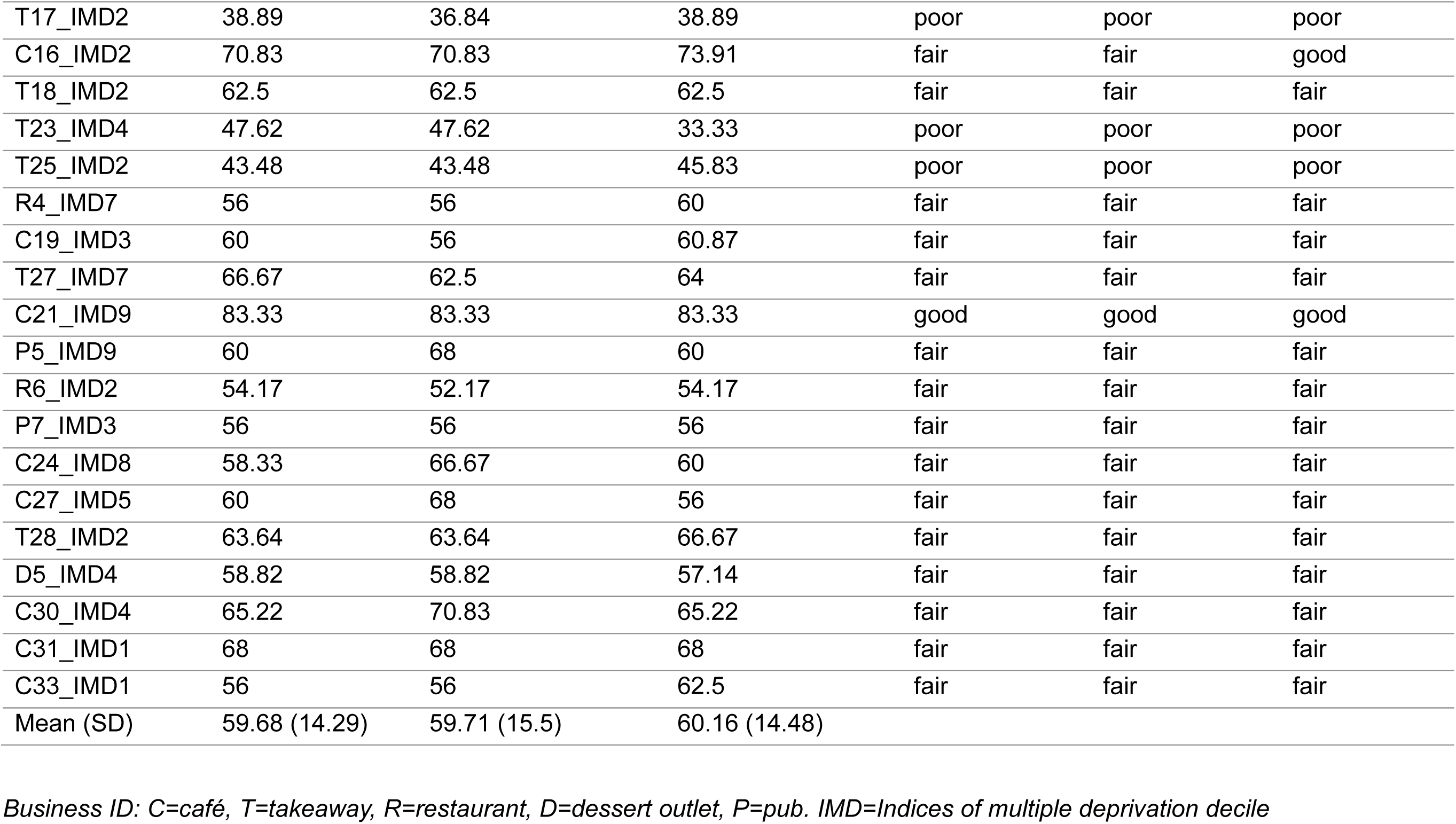
Individual rater and agreed healthiness scores and individual rater and agreed healthiness categories.

| Business ID | Agreed healthiness score (%) | Rater 1 healthiness score (%) | Rater 2 healthiness score (%) | Agreed healthiness category | Rater 1 healthiness category | Rater 2 healthiness category |
| --- | --- | --- | --- | --- | --- | --- |
| C1_IMD7 | 64 | 64 | 60 | fair | fair | fair |
| C5_IMD3 | 66.67 | 62.5 | 66.67 | fair | fair | fair |
| C8_IMD4 | 54.17 | 56 | 58.33 | fair | fair | fair |
| C10_IMD1 | 45.83 | 45.83 | 45.83 | poor | poor | poor |
| T4_IMD1 | 79.17 | 79.17 | 75 | good | good | good |
| D2_IMD1 | 25 | 12.5 | 25 | poor | poor | poor |
| C12_IMD1 | 61.9 | 47.83 | 68.18 | fair | poor | fair |
| T6_IMD1 | 63.64 | 63.64 | 63.64 | fair | fair | fair |
| C14_IMD2 | 60 | 60 | 60 | fair | fair | fair |
| T8_IMD1 | 39.13 | 39.13 | 39.13 | poor | poor | poor |
| T10_IMD1 | 43.48 | 43.48 | 39.13 | poor | poor | poor |
| T12_IMD1 | 47.83 | 47.83 | 52.17 | poor | poor | fair |
| T14_IMD1 | 43.48 | 43.48 | 43.48 | poor | poor | poor |
| T16_IMD1 | 35 | 33.33 | 35 | poor | poor | poor |
| C17_IMD3 | 68.18 | 68.18 | 85.71 | fair | fair | good |
| T19_IMD3 | 70.83 | 75 | 73.91 | fair | good | good |
| D4_IMD3 | 81.25 | 68.75 | 71.43 | good | fair | good |
| T20_IMD3 | 60 | 65 | 61.11 | fair | fair | fair |
| T22_IMD3 | 66.67 | 71.43 | 66.67 | fair | good | fair |
| T24_IMD3 | 73.91 | 73.91 | 68.18 | good | good | fair |
| R3_IMD6 | 77.27 | 86.36 | 68.75 | good | good | fair |
| C18_IMD3 | 66.67 | 66.67 | 66.67 | fair | fair | fair |
| T26_IMD7 | 63.64 | 68.18 | 80 | fair | fair | good |
| C20_IMD7 | 83.33 | 83.33 | 79.17 | good | good | good |
| P4_IMD7 | 68 | 69.23 | 75 | fair | fair | good |
| R5_IMD1 | 69.57 | 69.57 | 65.22 | fair | fair | fair |
| P6_IMD5 | 56.52 | 62.5 | 60.86 | fair | fair | fair |
| C22_IMD3 | 77.27 | 77.27 | 77.27 | good | good | good |
| C23_IMD10 | 65 | 65 | 66.67 | fair | fair | fair |
| C25_IMD5 | 68.18 | 70.83 | 69.57 | fair | fair | fair |
| C26_IMD1 | 69.57 | 69.57 | 69.57 | fair | fair | fair |
| C28_IMD2 | 75 | 75 | 75 | good | good | good |
| C32_IMD1 | 60 | 64 | 64 | fair | fair | fair |
| C2_IMD7 | 73.08 | 73.08 | 69.23 | good | good | fair |
| C6_IMD1 | 68 | 64 | 68 | fair | fair | fair |
| C7_IMD4 | 76 | 76 | 72 | good | good | good |
| C9_IMD3 | 72 | 72 | 72 | good | good | good |
| C11_IMD1 | 42.11 | 36.84 | 50 | poor | poor | fair |
| T5_IMD1 | 75 | 75 | 79.17 | good | good | good |
| D3_IMD2 | 12.5 | 12.5 | 12.5 | poor | poor | poor |
| C13_IMD2 | 46.15 | 46.67 | 41.67 | poor | poor | poor |
| T7_IMD3 | 43.48 | 43.48 | 45.83 | poor | poor | poor |
| C15_IMD4 | 70.83 | 70.83 | 66.67 | fair | fair | fair |
| T9_IMD4 | 34.78 | 34.78 | 34.78 | poor | poor | poor |
| T11_IMD2 | 54.55 | 54.55 | 57.14 | fair | fair | fair |
| T13_IMD2 | 38.89 | 33.33 | 44.44 | poor | poor | poor |
| T15_IMD1 | 52.17 | 52.17 | 52.17 | fair | fair | fair |
| T17_IMD2 | 38.89 | 36.84 | 38.89 | poor | poor | poor |
| C16_IMD2 | 70.83 | 70.83 | 73.91 | fair | fair | good |
| T18_IMD2 | 62.5 | 62.5 | 62.5 | fair | fair | fair |
| T23_IMD4 | 47.62 | 47.62 | 33.33 | poor | poor | poor |
| T25_IMD2 | 43.48 | 43.48 | 45.83 | poor | poor | poor |
| R4_IMD7 | 56 | 56 | 60 | fair | fair | fair |
| C19_IMD3 | 60 | 56 | 60.87 | fair | fair | fair |
| T27_IMD7 | 66.67 | 62.5 | 64 | fair | fair | fair |
| C21_IMD9 | 83.33 | 83.33 | 83.33 | good | good | good |
| P5_IMD9 | 60 | 68 | 60 | fair | fair | fair |
| R6_IMD2 | 54.17 | 52.17 | 54.17 | fair | fair | fair |
| P7_IMD3 | 56 | 56 | 56 | fair | fair | fair |
| C24_IMD8 | 58.33 | 66.67 | 60 | fair | fair | fair |
| C27_IMD5 | 60 | 68 | 56 | fair | fair | fair |
| T28_IMD2 | 63.64 | 63.64 | 66.67 | fair | fair | fair |
| D5_IMD4 | 58.82 | 58.82 | 57.14 | fair | fair | fair |
| C30_IMD4 | 65.22 | 70.83 | 65.22 | fair | fair | fair |
| C31_IMD1 | 68 | 68 | 68 | fair | fair | fair |
| C33_IMD1 | 56 | 56 | 62.5 | fair | fair | fair |
| Mean (SD) | 59.68 (14.29) | 59.71 (15.5) | 60.16 (14.48) |  |  |  |

*Business ID: C=café, T=takeaway, R=restaurant, D=dessert outlet, P=pub. IMD=Indices of multiple deprivation decile*

**Supplementary Material 5.** Inter-rater agreement and inter-rater reliability for individual tool criteria and tool categories

| Criterion number | Category | Inter-rater percent agreement | Inter-rater reliability: rater 1 and rater 2 score (Cohen's kappa) | Inter-rater reliability: rater 1, rater 2 and agreed score (Fleiss kappa) | Category percent agreement (Mean (SD)) | Category inter-rater reliability (Mean Cohen's kappa) | Category inter-rater reliability (Mean Fleiss kappa) |
| --- | --- | --- | --- | --- | --- | --- | --- |
| 1 | Healthier meal components | 98.48 | 0.93 | 0.95 | 94.55 (3.79) | 0.87 | 0.91 |
| 2 |  | 100 | 1 | 1 |  |  |  |
| 3 |  | 93.94 | 0.88 | 0.92 |  |  |  |
| 4 |  | 87.88 | 0.74 | 0.82 |  |  |  |
| 5 |  | 96.97 | 0.9 | 0.93 |  |  |  |
| 6 |  | 93.94 | 0.87 | 0.92 |  |  |  |
| 7 |  | 96.97 | 0.88 | 0.92 |  |  |  |
| 8 |  | 95.45 | 0.91 | 0.92 |  |  |  |
| 9 |  | 90.91 | 0.86 | 0.91 |  |  |  |
| 10 |  | 90.91 | 0.71 | 0.8 |  |  |  |
| 11 | Healthier drinks | 84.85 | 0.75 | 0.84 | 93.56 (6.35) | 0.84 | 0.90 |
| 12 |  | 100 | 1 | 1 |  |  |  |
| 13 |  | 93.94 | 0.69 | 0.8 |  |  |  |
| 14 |  | 95.45 | 0.92 | 0.95 |  |  |  |
| 15 | Healthier catering practices | 96.97 | 0.94 | 0.96 | 94.24 (2.71) | 0.86 | 0.90 |
| 16 |  | 93.94 | 0.81 | 0.87 |  |  |  |
| 17 |  | 92.42 | 0.89 | 0.92 |  |  |  |
| 18 |  | 96.97 | 0.93 | 0.95 |  |  |  |
| 19 |  | 90.91 | 0.73 | 0.81 |  |  |  |
| 20 | Portion size | 80.3 | 0.49 | 0.6 | 78.79 (2.14) | 0.52 | 0.64 |
| 21 |  | 77.27 | 0.54 | 0.68 |  |  |  |
| 22 | Special offers and promotions | 77.27 | 0.6 | 0.72 | 86.36 (7.88) | 0.46 | 0.63 |
| 23 |  | 90.91 | 0.42 | 0.59 |  |  |  |
| 24 |  | 90.91 | 0.36 | 0.57 |  |  |  |
| 25 | Diet and nutrition information | 90.91 | 0.76 | 0.84 | 95.46 (6.43) | 0.88 | 0.92 |
| 26 |  | 100 | 1 | 1 |  |  |  |

*Cohen's kappa interpretation: 0-0.20=no agreement, 0.21-0.39=minimal agreement, 0.40-0.59=weak agreement, 0.60-0.79=moderate* *agreement, 0.80-0.90=strong agreement and >0.90 almost perfect agreement [1]*
*Fleiss kappa interpretation: <0=poor agreement or no agreement, 0-0.20=slight agreement, 0.21-0.40=fair agreement, 0.41-0.60=moderate* *agreement, 0.61-0.80=substantial agreement, 0.81-1.00=almost perfect agreement [2]*

**Supplementary Material 6.**
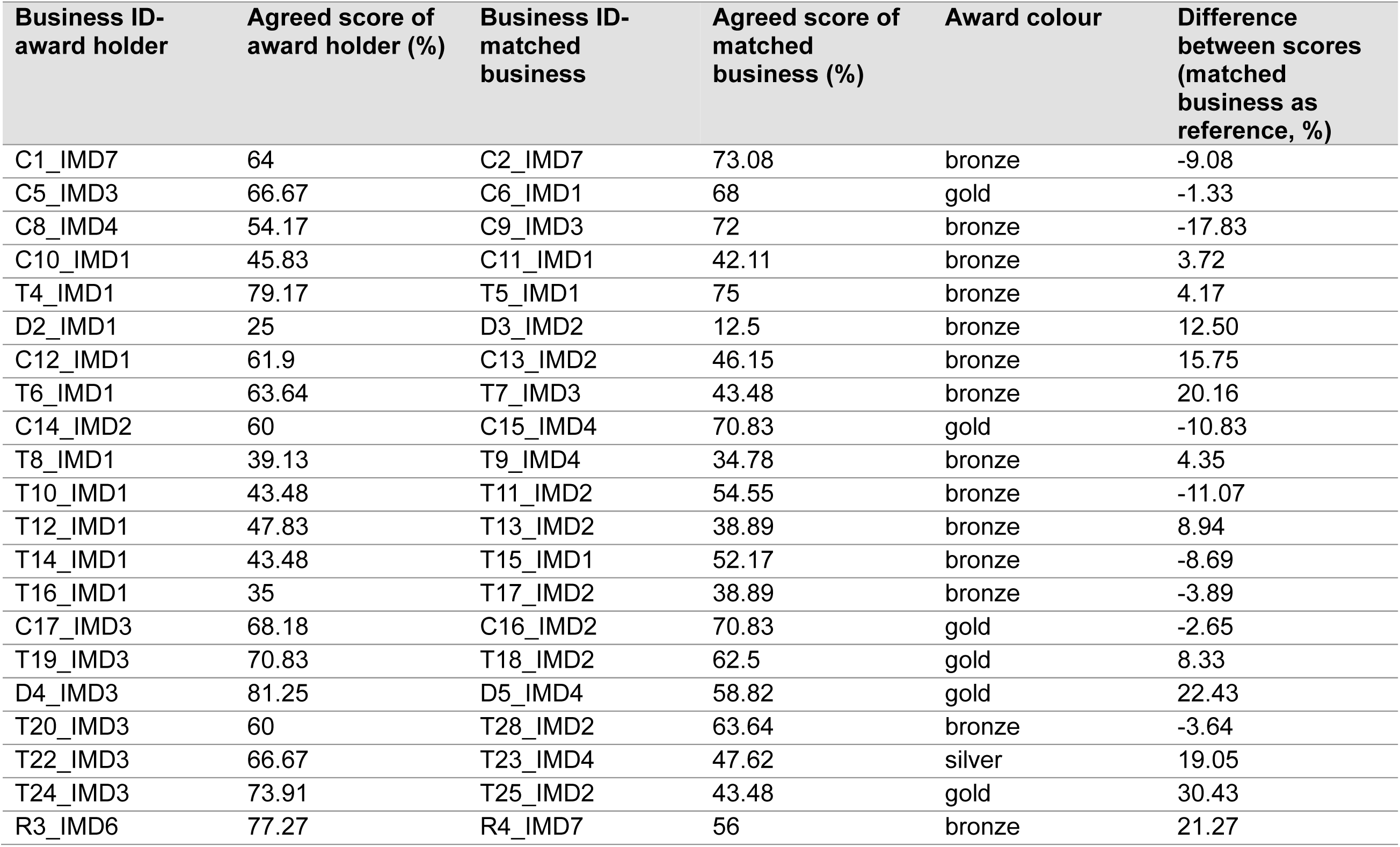
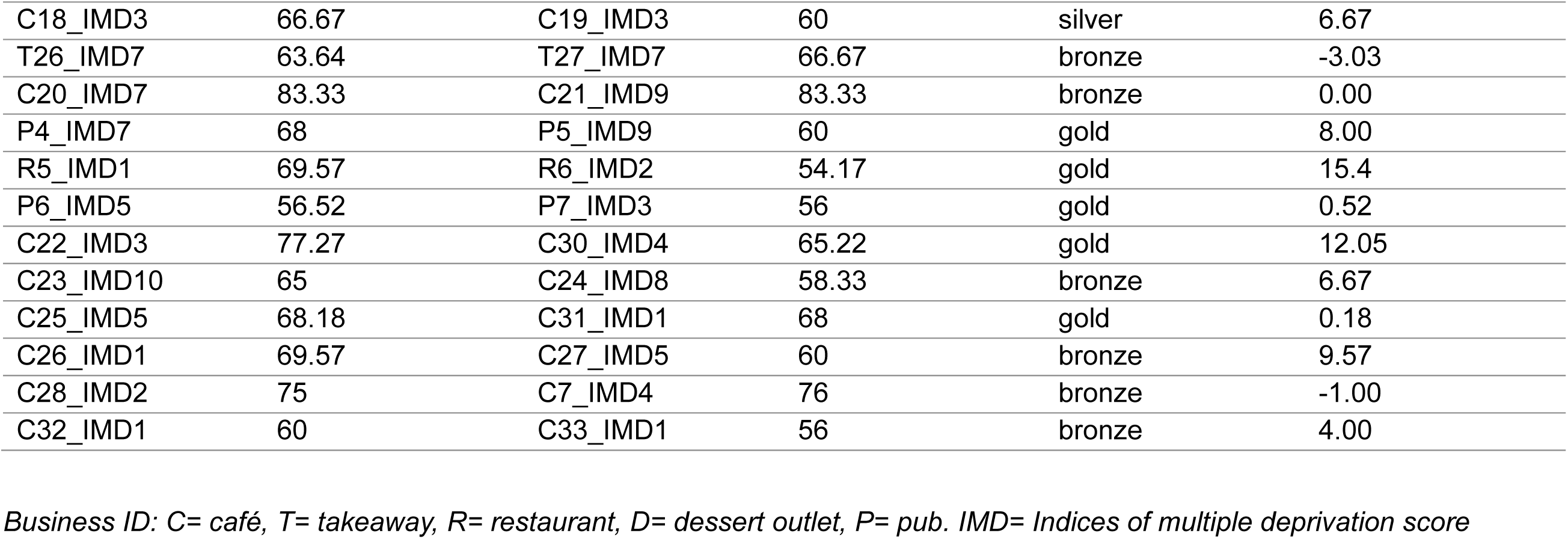
Comparison of agreed healthiness scores for businesses with the Recipe 4 health award vs. matched businesses without the award

Supplementary Material 7. Exploratory analyses: Comparison of healthiness scores with tool criteria removed

## Notes

### Competing Interest Statement

The authors have declared no competing interest.

### Clinical Protocols

https://osf.io/a743g/?view_only=a55924b192004570936f7bd385b1e902

### Funding Statement

This study was funded by the Economic and Social Research Council

### Author Declarations

Source data was publicly available through business websites or business visits

## References

[1] A. Blas, A. Garrido, O. Unver, and B. Willaarts, "A comparison of the Mediterranean diet and current food consumption patterns in Spain from a nutritional and water perspective," Science of The Total Environment, vol. 664, pp. 1020–1029, 2019/05/10/ 2019, doi: 10.1016/j.scitotenv.2019.02.111.

[2] A. Naska et al., "Eating out is different from eating at home among individuals who occasionally eat out. A cross-sectional study among middle-aged adults from eleven European countries," British Journal of Nutrition, vol. 113, no. 12, pp. 1951–1964, 2015, doi: 10.1017/S0007114515000963.

[3] E. Landais et al., "Consumption of food away from home in low- and middle-income countries: a systematic scoping review," Nutrition Reviews, vol. 81, no. 6, pp. 727–754, 2022, doi: 10.1093/nutrit/nuac085.

[4] H. G. Janssen, I. G. Davies, L. D. Richardson, and L. Stevenson, "Determinants of takeaway and fast food consumption: a narrative review," Nutrition Research Reviews, vol. 31, no. 1, pp. 16–34, 2018, doi: 10.1017/S0954422417000178.

[5] L. Biing-Hwan, J. Guthrie, and T. Smith, " Dietary Quality by Food Source and Demographics in the United States, 1977–2018. EIB-249,," U.S. Department of Agriculture, Economic Research Service, 2023. [Online]. Available: https://www.ers.usda.gov/publications/pub-details?pubid=105955

[6] S. J. Nielsen, A. M. Siega-Riz, and B. M. Popkin, "Trends in Energy Intake in U.S. between 1977 and 1996: Similar Shifts Seen across Age Groups," Obesity Research, vol. 10, no. 5, pp. 370–378, 2002, doi: 10.1038/oby.2002.51.

[7] J. Garbutt et al., "The contribution of the out-of-home food (OOHF) sector to the national diet: a cross-sectional survey with repeated 24-hour recalls of adults in England (2023- 2024)," medRxiv, p. 2025.06.30.25330369, 2025, doi: 10.1101/2025.06.30.25330369.

[8] Office for Health Improvements and Disparities, "National Diet and Nutrition Survey 2019 to 2023: report," 2025. [Online]. Available: https://www.gov.uk/government/statistics/national-diet-and-nutrition-survey-2019-to-2023/national-diet-and-nutrition-survey-2019-to-2023-report#food-and-drink-from-the-out-of-home-sector

[9] A. Jaworowska et al., "Nutritional composition of takeaway food in the UK," Nutrition & Food Science, vol. 44, no. 5, pp. 414–430, 2014, doi: 10.1108/nfs-08-2013-0093.

[10] A. Jaworowska, T. Blackham, and L. Stevenson, "Nutritional composition of takeaway meals served by independent small outlets," Proceedings of the Nutrition Society, vol. 70, no. OCE4, p. E166, 2011, Art no. E166, doi: 10.1017/S0029665111002175.

[11] L. E. Urban et al., "The Energy Content of Restaurant Foods Without Stated Calorie Information," JAMA Internal Medicine, vol. 173, no. 14, pp. 1292–1299, 2013, doi: 10.1001/jamainternmed.2013.6163.

[12] J. E. Todd, "Changes in consumption of food away from home and intakes of energy and other nutrients among US working-age adults, 2005–2014," *Public Health Nutrition*, vol. 20, no. 18, pp. 3238-3246, 2017, doi: 10.1017/S1368980017002403.

[13] J. Y. Polsky and D. Garriguet, "Eating away from home in Canada: impact on dietary intake," (in English), Health Reports, vol. 32, no. 8, pp. 26–34, Aug 2021 2021, doi: 10.25318/82-003-x202100800003-eng.

[14] R. A. Seguin, A. Aggarwal, F. Vermeylen, and A. Drewnowski, "Consumption Frequency of Foods Away from Home Linked with Higher Body Mass Index and Lower Fruit and Vegetable Intake among Adults: A Cross-Sectional Study," Journal of Environmental and Public Health, vol. 2016, no. 1, p. 3074241, 2016, doi: 10.1155/2016/3074241.

[15] Office for Health Improvements and Disparities. "Collection. Sugar, salt and calorie reduction and reformulation." https://www.gov.uk/government/collections/sugar-reduction#full-publication-update-history (accessed 29/10/2024).

[9] Public Health England. "Calorie reduction. Technical report: guidelines for industry, 2017 baseline calorie levels and the next steps." https://assets.publishing.service.gov.uk/media/5f560e4de90e0709942be6dd/Calorie_reduction_guidelines-Technical_report_070920-FINAL.pdf (accessed 29/10/2024).

[17] Department of Health and Social Care, "Press Release: New calorie labelling rules come into force to improve nation’s health," 2022. [Online]. Available: https://www.gov.uk/government/news/new-calorie-labelling-rules-come-into-force-to-improve-nations-health

[18] Department of Health and Social Care. "Calorie labelling in the out of home sector: implementation guidance " https://www.gov.uk/government/publications/calorie-labelling-in-the-out-of-home-sector/calorie-labelling-in-the-out-of-home-sector-implementation-guidance#introduction (accessed 29/10/2024).

[19] Department of Health and Social Care. "Government plans to tackle obesity in England. https://healthmedia.blog.gov.uk/2023/06/07/government-plans-to-tackle-obesity-in-england/." (accessed 03/01/2025).

[20] Z. Colombet, E. Robinson, C. Kypridemos, A. Jones, and M. O’Flaherty, "Effect of calorie labelling in the out-of-home food sector on adult obesity prevalence, cardiovascular mortality, and social inequalities in England: a modelling study," (in eng), Lancet Public Health, vol. 9, no. 3, pp. e178-e185, Mar 2024, doi: 10.1016/s2468-2667(23)00326-2.

[21] S. Bagwell, "Healthier catering initiatives in London, UK: an effective tool for encouraging healthier consumption behaviour?," Critical Public Health, vol. 24, no. 1, pp. 35–46, 2014/01/02 2014, doi: 10.1080/09581596.2013.769670.

[22] Public Health England. "Strategies for Encouraging Healthier ‘Out of Home’ Food Provision. A toolkit for local councils working with small food businesses." https://assets.publishing.service.gov.uk/media/5d83a91ee5274a27c5f4a8e8/Encouraging_healthier_out_of_home_food_provision_toolkit_for_local_councils.pdf (accessed 29/10/2024).

[23] Local Government Association. "Councillors’ guide to local authority public health responsibilities." https://www.local.gov.uk/publications/councillors-guide-local-authority-public-health-responsibilities-0 (accessed 16/06/2025).

[24] L. N. Gase, M. Kaur, L. Dunning, C. Montes, and T. Kuo, "What menu changes do restaurants make after joining a voluntary restaurant recognition program?," Appetite, vol. 89, pp. 131–135, 2015/06/01/ 2015, doi: 10.1016/j.appet.2015.01.026.

[25] J. J. Dwyer, L. A. Macaskill, C. L. Uetrecht, and C. Dombrow, "Eat Smart! Ontario’s Healthy Restaurant Program: focus groups with non-participating restaurant operators," (in eng), Can J Diet Pract Res, vol. 65, no. 1, pp. 6–9, Spring 2004, doi: 10.3148/65.1.2004.6.

[26] M. P. Fitzpatrick, G. E. Chapman, and S. I. Barr, "Lower-Fat Menu Items in Restaurants Satisfy Customers," Journal of the American Dietetic Association, vol. 97, no. 5, pp. 510- 514, 1997, doi: 10.1016/S0002-8223(97)00131-4.

[27] K. L. Green, S. L. Steer, R. E. Maluk, S. M. Mahaffey, and N. Muhajarine, "Evaluation of the Heart Smart Restaurant Program in Saskatoon and Regina, Saskatchewan," Canadian journal of public health, vol. 84, no. 6, pp. 399–402, 1993.

[28] C. D. Economos et al., "A community-based restaurant initiative to increase availability of healthy menu options in Somerville, Massachusetts: Shape Up Somerville," (in eng), Prev Chronic Dis, vol. 6, no. 3, p. A102, Jul 2009.

[29] A. H. Redelfs, J. D. Leos, H. Mata, S. L. Ruiz, and L. D. Whigham, "Eat Well El Paso!: Lessons Learned From a Community-Level Restaurant Initiative to Increase Availability of Healthy Options While Celebrating Local Cuisine," American Journal of Health Promotion, vol. 35, no. 6, pp. 841–844, 2021, doi: 10.1177/0890117121999184.

[30] M. Holdsworth, C. Haslam, and N. T. Raymond, "An assessment of compliance with nutrition criteria and food purchasing trends in Heartbeat Award premises," Journal of Human Nutrition and Dietetics, vol. 12, no. 4, pp. 327–335, 1999, doi: 10.1046/j.1365-277x.1999.00169.x.

[31] T. Brown, L. Vanderlinden, A. Birks, D. Mamatis, J. Levy, and T. Sahay, "Bringing Menu Labelling to Independent Restaurants: Findings from a Voluntary Pilot Project in Toronto," Canadian journal of dietetic practice and research, vol. 78, no. 4, pp. 177–181, 2017, doi: 10.3148/cjdpr-2017-014.

[32] E. T. Sosa, L. Biediger-Friedman, and M. Banda, "Associations Between a Voluntary Restaurant Menu Designation Initiative and Patron Purchasing Behavior," Health Promotion Practice, vol. 15, no. 2, pp. 281–287, 2014, doi: 10.1177/1524839912469535.

[33] A. Gupta et al., "Factors Influencing Implementation, Sustainability and Scalability of Healthy Food Retail Interventions: A Systematic Review of Reviews," Nutrients, vol. 14, no. 2, p. 294, 2022. [Online]. Available: https://www.mdpi.com/2072-6643/14/2/294.

[34] F. C. Hillier-Brown et al., "The impact of interventions to promote healthier ready-to-eat meals (to eat in, to take away or to be delivered) sold by specific food outlets open to the general public: a systematic review," (in eng), Obes Rev, vol. 18, no. 2, pp. 227–246, Feb 2017, doi: 10.1111/obr.12479.

[35] O. Huse et al., "The implementation and effectiveness of outlet-level healthy food and beverage accreditation schemes: A systematic review," (in eng), Obes Rev, vol. 24, no. 4, p. e13556, Apr 2023, doi: 10.1111/obr.13556.

[36] S. I. Kirkpatrick et al., "Dietary Assessment in Food Environment Research: A Systematic Review," American Journal of Preventive Medicine, vol. 46, no. 1, pp. 94–102, 2014/01/01/ 2014, doi: 10.1016/j.amepre.2013.08.015.

[37] B. Kelly, V. M. Flood, and H. Yeatman, "Measuring local food environments: An overview of available methods and measures," Health & Place, vol. 17, no. 6, pp. 1284–1293, 2011/11/01/ 2011, doi: 10.1016/j.healthplace.2011.08.014.

[38] B. E. Saelens, K. Glanz, J. F. Sallis, and L. D. Frank, "Nutrition Environment Measures Study in restaurants (NEMS-R): development and evaluation," (in eng), Am J Prev Med, vol. 32, no. 4, pp. 273–81, Apr 2007, doi: 10.1016/j.amepre.2006.12.022.

[39] K. Glanz et al., "Use of the Nutrition Environment Measures Survey: A Systematic Review," (in eng), Am J Prev Med, vol. 65, no. 1, pp. 131–142, Jul 2023, doi: 10.1016/j.amepre.2023.02.008.

[40] S. N. Partington, T. J. Menzies, T. A. Colburn, B. E. Saelens, and K. Glanz, "Reduced-Item Food Audits Based on the Nutrition Environment Measures Surveys," American Journal of Preventive Medicine, vol. 49, no. 4, pp. e23–e33, 2015/10/01/ 2015, doi: 10.1016/j.amepre.2015.04.036.

[41] J. E. Carins, S. Rundle-Thiele, and R. J. Storr, "Appraisal of short and long versions of the Nutrition Environment Measures Survey (NEMS-S and NEMS-R) in Australia," Public Health Nutrition, vol. 22, no. 3, pp. 564–570, 2019, doi: 10.1017/S1368980018002732.

[42] C. E. Pulker et al., "Development of the Menu Assessment Scoring Tool (MAST) to Assess the Nutritional Quality of Food Service Menus," (in eng), Int J Environ Res Public Health, vol. 20, no. 5, Feb 23 2023, doi: 10.3390/ijerph20053998.

[43] A. Rocha and C. Viegas, "KIMEHS—Proposal of an Index for Qualitative Evaluation of Children’s Menus—A Pilot Study," Foods, vol. 9, p. 1618, 11 2020, doi: 10.3390/foods9111618.

[44] L. F. Tavares et al., "Development and Application of Healthiness Indicators for Commercial Establishments That Sell Foods for Immediate Consumption," (in eng), Foods, vol. 10, no. 6, Jun 21 2021, doi: 10.3390/foods10061434.

[45] L. Goffe et al., "Supporting a Healthier Takeaway Meal Choice: Creating a Universal Health Rating for Online Takeaway Fast-Food Outlets.," International Journal of Environmental Research and Public Health., vol. 17(24):9260., 2020, doi: 10.3390/ijerph17249260.

[46] Action on Salt Action on Sugar. "Healthiness assessment in the UK out of home sector." https://www.actiononsalt.org.uk/media/action-on-salt/surveys/Healthiness-in-UK-OOH.pdf (accessed 16/06/2025).

[47] D. Cassady, R. Housemann, and C. Dagher, "Measuring Cues for Healthy Choices on Restaurant Menus: Development and Testing of a Measurement Instrument," American Journal of Health Promotion, vol. 18, no. 6, pp. 444–449, 2004, doi: 10.4278/0890-1171-18.6.444.

[48] S. S. Silva et al., "Development of a tool to assess the compliance of cafeteria menus with the Mediterranean Diet," *BMC Nutrition*, Article vol. 10, no. 1, 2024, Art no. 163, doi: 10.1186/s40795-024-00975-2.

[49] Public Health England. "Healthier Catering Guidance for Different Types of Businesses. Tips on providing and promoting healthier food and drink for children and families." https://assets.publishing.service.gov.uk/media/5d8330e9ed915d522c44704d/Healthier_catering_guidance_for_different_types_of_businesses.pdf (accessed 18/11/2024).

[50] Department for Environment Food and Rural Affairs. "Government Buying Standard for food and catering services." https://www.gov.uk/government/publications/sustainable-procurement-the-gbs-for-food-and-catering-services/government-buying-standard-for-food-and-catering-services#mandatory-standards (accessed 19/11/2024).

[51] House of Lords Food Diet and Obesity Committee. "Recipe for health: a plan to fix our broken food system." https://publications.parliament.uk/pa/ld5901/ldselect/ldmfdo/19/19.pdf (accessed 19/11/2024).

[52] National Health Service. "The Eatwell Guide." https://www.nhs.uk/live-well/eat-well/food-guidelines-and-food-labels/the-eatwell-guide/ (accessed 18/11/2024).

[53] Lancashire County Council. "Recipe 4 Health award." https://www.lancashire.gov.uk/business/trading-standards/recipe-4-health-award/ (accessed 06/11/2024).

[54] Food Active. "Pennine Lancashire Healthier Place Healthier Future Programme, ." https://foodactive.org.uk/what-we-do/campaigns-and-interventions/healthier-place-healthier-future/ (accessed 20/11/2024).

[55] Public Health England. "Composition of food integrated dataset (CoFID)." https://www.gov.uk/government/publications/composition-of-foods-integrated-dataset-cofid (accessed 24/07/2025).

[56] Department of Health, "Nutrient Profiling Technical Guidance ", 2011. [Online]. Available: https://assets.publishing.service.gov.uk/government/uploads/system/uploads/attachment_data/file/216094/dh_123492.pdf

[57] E. Gesteiro, A. García-Carro, R. Aparicio-Ugarriza, and M. González-Gross, "Eating out of Home: Influence on Nutrition, Health, and Policies: A Scoping Review," Nutrients, vol. 14, no. 6, p. 1265, 2022. [Online]. Available: https://www.mdpi.com/2072-6643/14/6/1265.

[58] L. Wellard-Cole, A. Davies, and M. Allman-Farinelli, "Contribution of foods prepared away from home to intakes of energy and nutrients of public health concern in adults: a systematic review," Critical Reviews in Food Science and Nutrition, vol. 62, no. 20, pp. 5511–5522, 2022/07/08 2022, doi: 10.1080/10408398.2021.1887075.

[59] M. L. McHugh, "Interrater reliability: the kappa statistic," Biochemia medica, vol. 22, no. 3, pp. 276–282, 2012, doi: 10.11613/bm.2012.031.

[60] E. Almanasreh, R. J. Moles, and T. F. Chen, "Chapter 41 - A practical approach to the assessment and quantification of content validity," in *Contemporary Research Methods in Pharmacy and Health Services*, S. P. Desselle, V. García-Cárdenas, C. Anderson, P. Aslani, A. M. H. Chen, and T. F. Chen Eds.: Academic Press, 2022, pp. 583-599.

[61] World Health Organization. "Alcohol Fact Sheet." https://www.who.int/news-room/fact-sheets/detail/alcohol (accessed 02/07/2025).

[62] D. F. Polit and C. T. Beck, "The content validity index: are you sure you know what’s being reported? Critique and recommendations," (in eng), Res Nurs Health, vol. 29, no. 5, pp. 489–97, Oct 2006, doi: 10.1002/nur.20147.

[63] A. K. Taher, N. Evans, and C. E. L. Evans, "The cross-sectional relationships between consumption of takeaway food, eating meals outside the home and diet quality in British adolescents," Public Health Nutrition, vol. 22, no. 1, pp. 63–73, 2019, doi: 10.1017/S1368980018002690.

[64] C. Jostock, H. Forde, N. Roberts, S. A. Jebb, R. Pechey, and L. Bandy, "Healthy eating interventions conducted in small, local restaurants and hot food takeaways: a systematic review," Public Health Nutrition, vol. 28, no. 1, p. e24, 2025, Art no. e24, doi: 10.1017/S1368980025000035.

[65] E. R. Maguire, T. Burgoine, and P. Monsivais, "Area deprivation and the food environment over time: A repeated cross-sectional study on takeaway outlet density and supermarket presence in Norfolk, UK, 1990–2008," *Health & Place*, vol. 33, pp. 142-147, 2015/05/01/ 2015, doi: 10.1016/j.healthplace.2015.02.012.

[66] Y. Huang, T. Burgoine, T. R. P. Bishop, and J. Adams, "Assessing the healthiness of menus of all out-of-home food outlets and its socioeconomic patterns in Great Britain," Health & Place, vol. 85, p. 103146, 2024/01/01/ 2024, doi: 10.1016/j.healthplace.2023.103146.

[67] Food Standards Agency. "Food hygiene ratings." https://ratings.food.gov.uk/ (accessed 19/11/2024).

[68] G. Rau and Y.-S. Shih, "Evaluation of Cohen’s kappa and other measures of inter-rater agreement for genre analysis and other nominal data," Journal of English for Academic Purposes, vol. 53, p. 101026, 2021/09/01/ 2021, doi: 10.1016/j.jeap.2021.101026.

[69] N. Gisev, J. S. Bell, and T. F. Chen, "Interrater agreement and interrater reliability: Key concepts, approaches, and applications," Research in Social and Administrative Pharmacy, vol. 9, no. 3, pp. 330–338, 2013/05/01/ 2013, doi: 10.1016/j.sapharm.2012.04.004.

[70] S. Kılıç, "Kappa testi," Journal of mood disorders, vol. 5, no. 3, pp. 142–144, 2015.

[71] J. R. Landis and G. G. Koch, "The Measurement of Observer Agreement for Categorical Data," Biometrics, vol. 33, no. 1, pp. 159–174, 1977, doi: 10.2307/2529310.

[72] T. K. Koo and M. Y. Li, "A Guideline of Selecting and Reporting Intraclass Correlation Coefficients for Reliability Research," (in eng), J Chiropr Med, vol. 15, no. 2, pp. 155–63, Jun 2016, doi: 10.1016/j.jcm.2016.02.012.

[73] R Core Team, "R: A language and environment for statistical computing. R Foundation for Statistical Computing, Vienna, Austria.," 2021. [Online]. Available: URL https://www.R-project.org/.

[74] J. Cohen, "A Coefficient of Agreement for Nominal Scales," Educational and Psychological Measurement, vol. 20, no. 1, pp. 37–46, 1960, doi: 10.1177/001316446002000104.

[75] K. E. Smoyer-Tomic et al., "The association between neighborhood socioeconomic status and exposure to supermarkets and fast food outlets," Health & Place, vol. 14, no. 4, pp. 740–754, 2008/12/01/ 2008, doi: 10.1016/j.healthplace.2007.12.001.

[76] L. A. Lytle and R. L. Sokol, "Measures of the food environment: A systematic review of the field, 2007–2015," *Health & Place*, vol. 44, pp. 18-34, 2017/03/01/ 2017, doi: 10.1016/j.healthplace.2016.12.007.

[77] L. Rimkus et al., "Development and Reliability Testing of a Fast-Food Restaurant Observation Form," American Journal of Health Promotion, vol. 30, no. 1, pp. 9–18, 2015, doi: 10.4278/ajhp.130731-QUAN-389.

[78] Office for Health Improvements and Disparities, "Obesity Profile: short statistical commentary May 2024," 03/02/2025 2024, doi: https://www.gov.uk/government/statistics/update-to-the-obesity-profile-on-fingertips/obesity-profile-short-statistical-commentary-may-2024.

[79] M. A. Nagi et al., "Economic costs of obesity: a systematic review," International Journal of Obesity, vol. 48, no. 1, pp. 33–43, 2024/01/01 2024, doi: 10.1038/s41366-023-01398-y.

## References

[1] M. L. McHugh, "Interrater reliability: the kappa statistic," Biochemia medica, vol. 22, no. 3, pp. 276–282, 2012, doi: 10.11613/bm.2012.031.

[2] J. R. Landis and G. G. Koch, "The Measurement of Observer Agreement for Categorical Data," Biometrics, vol. 33, no. 1, pp. 159–174, 1977, doi: 10.2307/2529310.

